# Lessons learnt from the use of compartmental models over the COVID-19 induced lockdown in France

**DOI:** 10.1101/2021.01.11.21249565

**Authors:** Romain Gauchon, Nicolas Ponthus, Catherine Pothier, Christophe Rigotti, Vitaly Volpert, Stéphane Derrode, Jean-Pierre Bertoglio, Alexis Bienvenüe, Pierre-Olivier Goffard, Anne Eyraud-Loisel, Simon Pageaud, Jean Iwaz, Stéphane Loisel, Pascal Roy

**Author notes:** Equal contributor, order is alphabetical (Romain Gauchon, Nicolas Ponthus). For the group CovDyn (Covid Dynamics). Members of the group are listed at the end of the document.

## Abstract

**Background:** Compartmental models help making public health decisions. They were used during the COVID-19 outbreak to estimate the reproduction numbers and predict the number of hospital beds required. This study examined the ability of closely related compartmental models to reflect equivalent epidemic dynamics.

**Methods:** The study considered three independently designed compartmental models that described the COVID-19 outbreak in France. Model compartments and parameters were expressed in a common framework and models were calibrated using the same hospitalization data from two official public databases. The calibration procedure was repeated over three different periods to compare model abilities to: i) fit over the whole lockdown; ii) predict the course of the epidemic during the lockdown; and, iii) provide profiles to predict hospitalization prevalence after lockdown. The study considered national and regional coverages.

**Results:** The three models were all flexible enough to match real hospitalization data during the lockdown, but the numbers of cases in the other compartments differed. The three models failed to predict reliably the number of hospitalizations after the fitting periods at national as at regional scales. At the national scale, an improved calibration led to epidemic course profiles that reflected hospitalization dynamics and reproduction numbers that were coherent with official and literature estimates.

**Conclusion:** This study shows that prevalence data are needed to further refine the calibration and make a selection between still divergent models. This underlines strongly the need for repeated prevalence studies on representative population samples.

## 1 Background

The COVID-19 coronavirus outbreak is causing a major national, European, and international health crisis, with serious consequences in terms of public health. The COVID-19 epidemic started in China in late autumn 2019 and on December 31, the WHO China Country Office [1] received reports of severe pneumonia cases in the city of Wuhan (Hubei Province). The magnitude of the epidemic (currently more than one million officially recognized deaths worldwide) has led to exceptional reactions from different countries around the world. Due to the early outbreak in Italy, Europe has been one of the most severely hit continents. According to worldometers, the continent (including Russia) counts now more than 290 000 coronavirus-related deaths. On March 9, 2020, the Italian government started a national lockdown and most European countries followed, albeit with some variations between countries.

In France, the lockdown began officially on March 17 at noon, although some measures, such as restaurant closure, occurred days before that date. During the lockdown, individuals could leave home only for individual physical activity, work, essential goods, or medical reasons. Thus, the social behavior changed drastically resulting in a drop of *R*_0_ coefficient (i.e., the average number of people infected by a single infected person). As the national situation improved, the lockdown was lifted on May 12, resulting in a change in the contact matrix.The unprecedented implementation of a lockdown system stimulated research in epidemics modeling. Epidemiologists investigated the impact of the lockdown in France (e.g., see [2, 3, 4]) through compartmental models. The main goal was to provide public authorities decision-making tools. Assessing the way these models perform regarding the COVID-19 pandemic and differences between them is essential to check their reliability and usefulness in case of future waves.

One peculiarity of COVID-19 outbreak is the limited amount of data available for epidemic modeling. This is due to many factors such as the asymptomatic form of the disease that represents a non-negligible percentage of cases. This motivated a number of analyses aimed at predicting the consequences of the pandemic on hospital activities (occupation of hospital and intensive care unit beds) and in terms of COVID-19-linked hospital death toll. Compartmental models were then fitted using only hospital source data and other literature parameters to calibrate non-observed compartments.

This study aims to assess the statistical properties of three different compartmental models designed to describe the COVID-19 outbreak in France. These models were proposed by research teams from University of Bordeaux (Prague et al. [5]), INSERM (Di Domenico et al. [6]), and EHESP (Roux et al. [7]). The abilities of these models to describe the dynamics of the epidemic before and during lockdown (phases I and II) and predict these dynamics after the lockdown (phase III) according to each of the three models under several hypotheses. Because these models have major structural differences, the study compared also the impacts of these differences on the epidemic dynamics.

## 2 Data and methods

### 2.1 Data

Two different French public databases were used. The main one was the database of Santé Publique France (SPF, available on data.gouv.fr). Since March 18, this database provides aggregate information on a day-to-day basis about every patient hospitalized in a private or public hospital, including age (or age group), sex and, most importantly, the daily number of new COVID-19 cases admitted and number of occupied hospital beds. This SPF’s database includes no data previous to March 18. Thus, data from the “Surveillance sanitaire des urgences et des décès” (SurSaUD) database were used as complement. SurSaUD is a French health monitoring system that started recording information on patients with COVID-19 symptoms who visited hospital emergency units since February 24. However, this database covers only 86% of emergency health units. This underestimation was corrected using the process proposed by Salje et al. [2], based on the daily ratio of SPF to SurSaUD number of cases. This ratio was calculated between March 18 and March 27, and the median of these values was then used to estimate and correct SurSaUD data. The SurSaUD database was used to estimate the number of patients hospitalized on March 2 (starting date of modeling) as well as the dynamics of the epidemic from March 8 to March 18 (i.e., phase I, before lockdown).

### 2.2 Statistical modelling

#### 2.2.1 Adapted models

##### Modeling the disease contagiousness

To take into account possible differences in contagiousness between young and old people, the population was divided into *n* age groups. For a given age group *i*, at a given time *t*, the incidence hazard rate 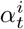 was calculated as follows:

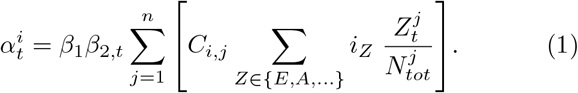

In this equation,

- *C* is the contact matrix within which *C*_*i,j*_ is the average number of individuals from age group *j* encountered per day by a single individual from age group *i*;
- *Z* denotes a compartment in the set of all compartments, *Z* ∈ {*E*, *A*, …} (see Figure 1 for the compartment names in the three models);
- 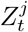 is the number of individuals of age group *j* in compartment *Z* at time *t*;
- 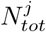 is the total number of individuals from age group *j* (who contracted the disease or not);
- Thus, the term into brackets is the daily average number of infected individuals of age group *j* who came into contact with one individual of age group *i*;
- *i*_*Z*_ is the relative infectivity of individuals in compartment *Z*. When individuals in compartment *Z* are not contagious (e.g., if they have not contracted the disease yet), then *i*_*Z*_ = 0. Some compartments, such as the one relative to asymptomatic subjects, could have a relative infectivity ranging between 0 and 1;
- *β*_1_ is the daily incidence hazard rate in case of one daily contact with a contagious person (the parameter is estimated for a standard duration contact);
- The relative hazard *β*_2,*t*_ estimates the effect of lockdown by reducing the mean number of daily contacts, and/or the mean duration of contacts, and/or their infectivity under barrier measures.

**Figure 1:**
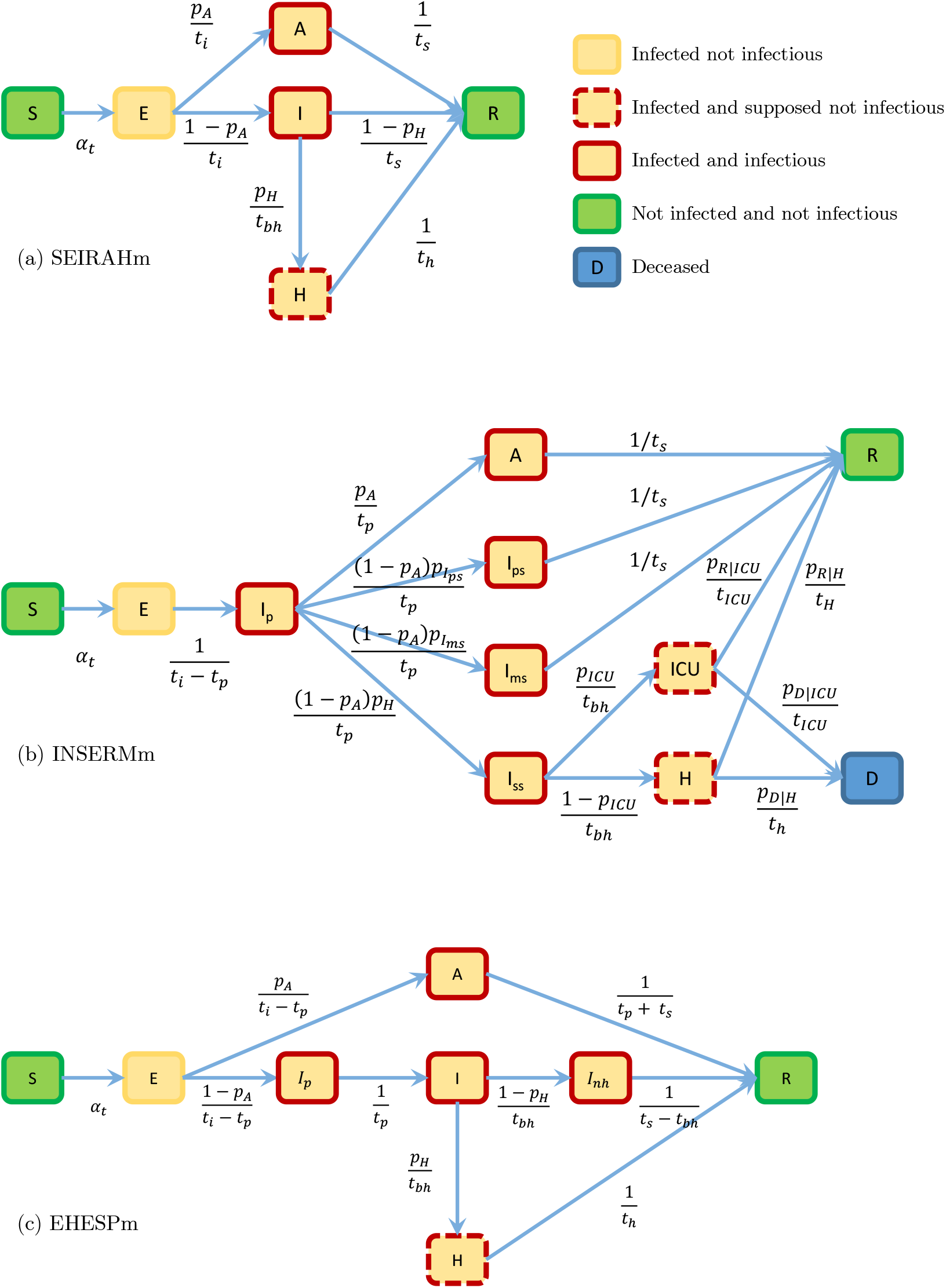
Diagrams of the models.

In the case of a single age group, Equation 1 becomes:

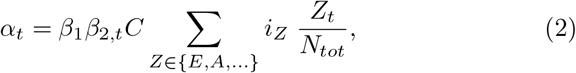

where *C* is the mean number of daily contacts.

##### Lockdown

In France, the lockdown started on March 17 at noon and ended on May 10 at midnight. During this period, individuals could go out to work, buy essential goods, seek medical care, or practice an individual sports activity. The daily incidence hazard rate decreased (*β*_2,*t*_ *<* 1, see Equation (1)) leading to a drop in the basic reproduction number *R*_0_. Given a starting date of the model at *t* = 0, let *T*_*b*_ and *T*_*e*_ be the times of the beginning^[1]^ and end of the lockdown respectively. After the beginning of the lockdown, *β*_2,*t*_ was assumed to change linearly^[2]^ over time until reaching a plateau at time *T*_*c*_.

After the end of the lockdown, *β*_2,*t*_ was assumed to change linearly again, until reaching a new plateau at time *T*_*p*_. The relative hazard *β*_2,*t*_ verifies

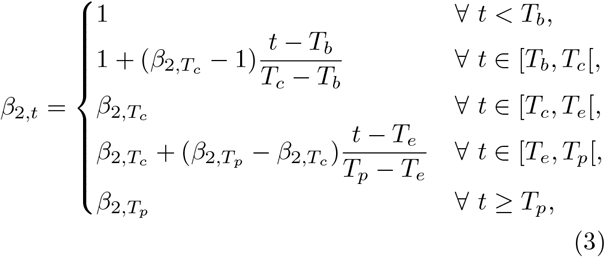

with *T*_*b*_ *≤ T*_*c*_ *≤ T*_*e*_ *≤ T*_*p*_. In this framework, *T*_*c*_ and 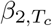 had to be estimated^[3]^. A sudden fall of *β*_2,*t*_ would have led to a discontinuity in the daily infection rate and, consequently, to a discontinuity in the number of daily hospitalizations. Such a discontinuity was not observed. The smooth change in the number of daily hospitalizations seemed related to a continuous change in *β*_2_ and, consequently, *R*_0_. Term 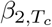 is allowed to vary with the area modeled.

##### SEIRAH model

This model (herein noted SEIRAHm) was adapted from Prague et al. [5]. This model was the simplest of all three models considered because it did not consider age. Thus, no contact matrix was considered.

The model consisted of six compartments:

- *S* (Susceptible): individuals who were not infected by the virus;
- *E* (Exposed): individuals who contracted the virus. This compartment corresponds to disease incubation and individuals were supposed not to be infectious. Individuals in this class become either asymptomatic virus carriers (with probability *p*_*A*_), or symptomatic carriers (with probability 1 − *p*_*A*_);
- (Infected): individuals who presented disease symptoms without being hospitalized. These individuals become either hospitalized patients (with probability *p*_*H*_), or Removed (with probability 1 − *p*_*H*_);
- *R* (Removed): this compartment groups together individuals cured or deceased from the disease;
- *A* (Asymptomatic): individuals who completed the incubation period, became infectious, but had no disease symptoms. Unlike population in compartment *I*, this population had no cough, fever, etc. The relative infectivity of these asymptomatic individuals was estimated at *i*_*A*_;
- *H* (Hospitalized): hospitalized individuals, (ICU or else). Since COVID-19 patients were usually isolated at hospital admission, their infectivity was fixed at zero though this was not entirely true (they could infect their caregivers).

A synthetic view of this model is shown in Figure 1a, and the parameters are summarized in Appendix A.2. The dynamics of the model was given by Equation system (4).

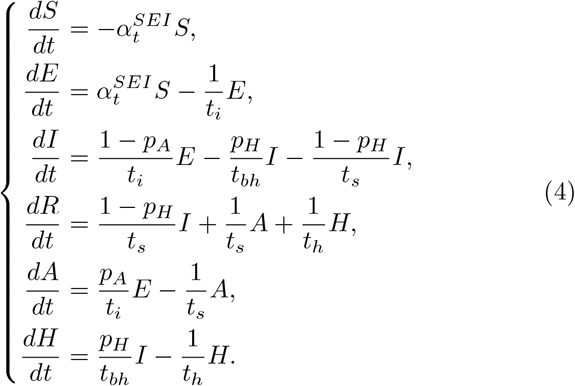

This model has two differences with the original model of Prague et al.: i) the addition of a probability for hospitalization; and, ii) *t*_*bh*_ was not estimated but fixed at a value taken from [2]. Not estimating *t*_*bh*_ allowed a fair comparison with INSERMm and EHESPm. Thus, the three models had the same number of inferred parameters (see Appendix A.3). A compartment between *I* and *R* could be created to allow for an average time spent by nonhospitalized symptomatic individuals in compartment *I* (similar to the compartment *I*_*nh*_ in EHESPm), but this was not done to keep the model close to the original one. Parameter 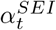 was obtained from Equation (2).

Here, according to [8] the average daily number of contact *C* in France was equal to 12. Originally, in [5], compartment *I* was supposed to include individuals tested positive and compartment *A* to include all other infected but not tested cases.

This model was originally fitted to both incidence in *H* and incidence of new tested cases in *I* using Poisson distributions to model the likelihood.

##### INSERM model

This model (herein noted INSERMm) was adapted from the model described in [6]. It is a compartmental model stratified by age groups. For each age group, the structure shown in Figure 1b leads to the following system of differential equations:

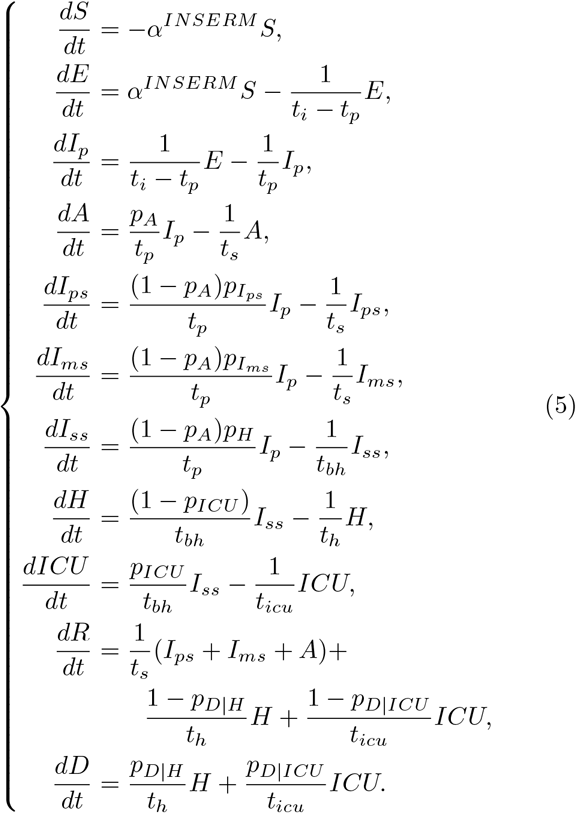

The *α*^*INSERM*^ parameter was obtained from Equation (1). The compartments of this model and the differences with SEIRAHm are the following:

- *S* (Susceptible): individuals who were not infected by the virus (same as *S* in SEIRAHm);
- Unlike the SEIRAHm, the incubation period included two contiguous compartments:
  – *E* (Exposed): infected individuals who did not develop symptoms and were not contagious;
  – *I*_*p*_ (Prodromic^[4]^ phase): individuals still in the incubation period without apparent symptoms but already contagious.
  The average time spent in compartment *E* in SEIRAHm is thus equal to the sum of the average times spent in compartments *E* and *I*_*p*_ of INSERMm^[5]^;
- The symptomatic infectious period included three compartments:
  – *I*_*ps*_ (Paucisymptomatic disease): individuals with weak disease symptoms;
  – *I*_*ms*_ (Medium symptoms): individuals with disease symptoms (e.g., fever or cough) who did not require hospitalization;
  – *I*_*ss*_ (Severe symptoms): individuals severely infected who required hospitalization; the probability of going into this compartment was equal to *p*_*H*_.
  Compartment *I* of SEIRAHm was thus analogous to the sum of compartments *I*_*ps*_, *I*_*ms*_ and *I*_*ss*_ of INSERMm.
- In this model, there are two absorbing compartments:
  – *D* (Deceased at hospital): This compartment had no equivalent in SEIRAHm. The benefit from considering only hospital deaths was motivated by the availability of these data;
  – *R* (Removed): individuals who recovered or died from the disease out of hospital.
  Compartment *R* of SEIRAHm is thus analogous to the sum of compartments *R* and *D* of INSERMm;
- *A* (Asymptomatic): individuals who completed the incubation period, became infectious, but had no disease symptoms (same as in SEIRAHm);
- The hospitalization period included two compartments:
  – *ICU* (Intensive Care Units): individuals treated at any time in an ICU; here, it includes both pre- and post-ICU periods;
  – *H* (Hospitalized): individuals hospitalized without requiring ICU care.
  Compartment *H* of SEIRAHm is analogous to the sum of compartments *H* and *ICU* of INSERMm.

INSERMm was originally developed with three age groups [6]. To obtain results easy to compare with those of EHESPm, it was implemented with 17 age groups. To allow comparisons between models, all common parameters were defined the same way. This implied some changes to the original model [6] (e.g., to obtain one parameter for each of the 17 age groups instead of 3 in the original model). In particular:

- In INSERMm [6], the average times spent in compartments *A, I*_*ps*_, *I*_*ms*_, and *I*_*ss*_ were considered equal. Here, the average time for compartment *I*_*ss*_ was set to *t*_*bh*_ denoting the time before hospitalization.
- The allocation of infected individuals between compartments *I*_*ps*_ and *I*_*ms*_ was not used when comparing this model results to those of other models. Therefore, *p*_*I*_*ps* and *p*_*I*_*ms* were arbitrarily and respectively fixed to one quarter and three quarters of (1 − *p*_*H*_);
- The original model [6] used different hazard rates for transitions from compartments *H* and *ICU* to compartments *R* and *D*. Here, only two average times were used (*t*_*h*_ and *t*_*icu*_).

The Next Generation Matrix approach proposed by Diekmann et al. [9] was applied to calculate the basic reproduction number for this model (for more details, see Appendix A.4).

Originally, in [6], this model was fitted to the incidence in compartment *H* using a Poisson distribution to model the likelihood.

##### EHESP model

This model (herein noted EHESPm) was proposed by Roux et al. [7]. It is described here by Figure 1c and Equation system (6).

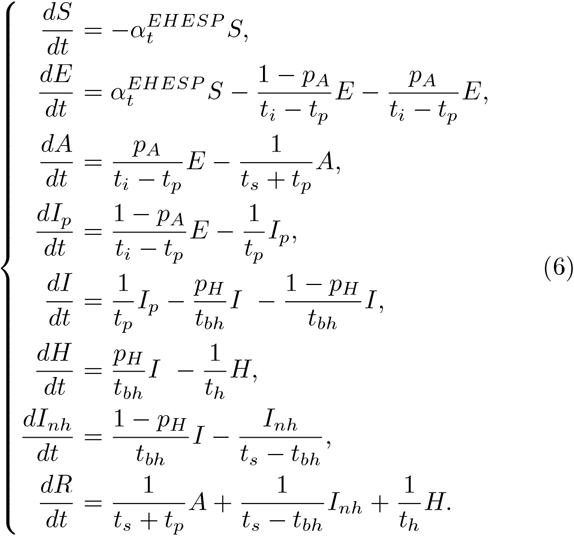

The 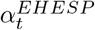 parameter is given by Equation (1). As INSERMm, EHESPm may be considered as a sophisticated version of SEIRAHm. The compartments and the differences are the following:

- *S* (Susceptible): individuals who were not infected by the virus (same as in SEIRAHm);
- The incubation period included two compartments as in INSERMm but contrary to SEIRAHm:
  – *E* (Exposed): infected individuals who did not develop symptoms and were not contagious;
  – *I*_*p*_ (Prodromic phase): individuals still in the incubation period without apparent symptoms but already contagious. The average time spent in compartment *E* of SEIRAHm was thus equal to the sum of the average times spent in compartments *E* and *I*_*p*_ in EHESPm;
- The symptomatic infectious phase included two compartments:
  – *I* (Infected): symptomatic individuals still at an early stage of the disease;
  – *I*_*nh*_ (Infected non-hospitalized): symptomatic individuals who did not need hospitalization;
  Compartment *I* of SEIRAHm was thus analogous to the sum of compartments *I* and *I*_*nh*_ of EHESPm. However, there was a small but interesting difference between the two models: SEIRAHm supposed implicitly that an individual could go to hospital at any time, whereas EHESPm supposed implicitly that when an individual has been ill for some (long) time, he/she would not go to hospital, even in case of worsening of symptoms (he/she would then stay artificially in compartment *I*_*nh*_);
- *R* (Removed): groups together individuals cured or died from the disease (same as in SEIRAHm);
- *A* (Asymptomatic): individuals who completed the incubation period, became infectious, but had no disease symptoms (same as in SEIRAHm);
- *H* (Hospitalized): same compartment as in SEIRAHm.

Originally, in [7], this model was fitted on both incidence and prevalence data of compartment *H* using respectively a negative binomial distribution and a Poisson distribution to model the likelihood.

##### 2.2.2 Supplementary material

Other technical details are presented in Appendix A, which is organized as follows.

- Appendix A.1 details the initialization process. In this process, only one compartment by model was automatically estimated during the calibration process: the number of individuals at date *t* = 0 in compartment *E*. Other compartments were initialized using SurSaUD data and exploiting the properties of the models. All models were assumed to start on March 2, 2020.
- Appendix A.2 summarizes the notations, values, and references of the parameters taken from the literature.
- Appendix A.3 describes the estimation process used for model calibration.
- Appendix A.4 details the method used to compute *R*_0_ and *R*_*e*_. The theory is recalled and illustrated with the three models.

##### 2.2.3 The modelling strategy

The modelling strategy is organized around three main research axes presented respectively in the next three paragraphs. A unified simple approach based on the maximum likelihood was applied to the three models to fit the changes in prevalent hospitalized cases.

The respective abilities of the three models to describe the dynamics of the epidemic during the whole lockdown period was analyzed by comparing the observed numbers of hospitalized patients with the numbers in compartment *H* of SEIRAHm and EHESPm as well as with the sum in compartments*H* and *ICU* of INSERMm. The maximum likelihood estimates of the parameters of the three models were obtained by fitting the daily numbers of prevalent hospitalized cases recorded in SPF database during the whole lockdown period (from March 18 to May 11, 2020, F-WL approach: fit over the whole lockdown). The predicted daily incident hospitalized cases (not used to fit the models) were graphically compared with the observed ones. Although the corresponding observed numbers were not available, the predicted daily numbers of patients in compartments *E, I* and *R* were compared between the three models. Moreover, the French government published daily the number of beds occupied by COVID-19 patients and the number of COVID-19 deaths at hospital. Because INSERMm has *ICU* and *D* compartments, joint changes of fitted vs. observed numbers in these two compartments were compared in this model only.

The abilities of the three models to predict the course of the epidemic during the lockdown period was analyzed by fitting the models on the numbers of hospitalized patients in the SPF database at the start of the lockdown (March 18 to April 6, F-SL approach: fit at start of lockdown). The predictions of the models for period April 7 to May 11 were then compared to the observed data.

The dynamics of the epidemic as predicted by the three models after the lockdown were compared to the observed data for different values of *R*_0_. Parameter *β*_2,*t*_ was supposed to change linearly from May 11 to June 2. The study computed with each model the values of *β*_2,*t*_ that correspond to 15 values of *R*_0_ ranging from 0.1 to 1.5 by 0.1 increments as well as the corresponding profile likelihood. The predicted change in the *R*_0_ that fitted the best the observed data over a one-month period (May 11 to June 10) according to the maximum likelihood was then retained. Two approaches were compared. In the first, model parameters were estimated by fitting all lockdown data (March 18 to May 11, F-WL approach). In the second, parameters *E*_0_ and *β*_1_ were estimated using the corrected SurSaUD database from March 8 to March 18. The objective was to estimate the parameters not linked to the lockdown with data previous to the lockdown. Then, the last two parameters (*T*_*c*_ and 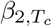, from Equation (3)) were estimated using SPF data from May 1 to May 11, which were almost not influenced by the number of individuals infected before the lockdown and thus gave more accurate estimates of the parameters resulting from the lockdown. This fit of the epidemic process restricted to the end of the lockdown was intended to provide more accurate estimates of 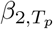. Estimates of *R*_0_ and *R*_*e*_ with the second approach (herein F-ALEL: fit ante lockdown and end of lockdown) at the end of the lockdown were computed (as described in Appendix A.4) and compared with those found in the literature.

The three main axes were illustrated for whole France and four regions: Auvergne-Rhône-Alpes (ARA), Île de France (IDF), Grand Est (GE), and Nouvelle Aquitaine, Occitanie, and Provence-Alpes Côte d’Azur (SUD).

## 3 Results

The mains results for whole France are presented herein, whereas complementary results are available in Appendix B.

In each Figure, the vertical red dotted line represents the date of lockdown and the vertical blue dotted line the limit dates used for parameter calibration when they differed from the lockdown dates.

### 3.1 Description of the epidemic during the whole lockdown period

At the national level, the fit of the three models on case prevalence data in the compartment *H* (*H* + *ICU* in INSERMm) was good (Figure 2a). The three models described quite accurately the daily numbers of individuals in compartment *H*. However, the adequacy between the daily number of individuals arriving to compartment *H* and the observed data published by SPF differed in various ways depending on the model considered (Figure 2b). With SEIRAHm, the estimated daily number of individuals arriving to compartment *H* reached its maximum before (and at a lower level) the two other models. The results with SEIRAHm were less accurate at mid-lockdown but more accurate at end of lockdown, whereas EHESPm and INSERMm overestimated the incidence of hospitalization. Parameters *E*_0_ and *β*_2_, as estimated by SEIRAHm model, were smaller than those estimated by EHESPm or INSERMm (Table B.1 in Appendix B.1). On the contrary, parameter *β*_1_ was higher with SEIRAHm than with the two other models.

**Figure 2:**
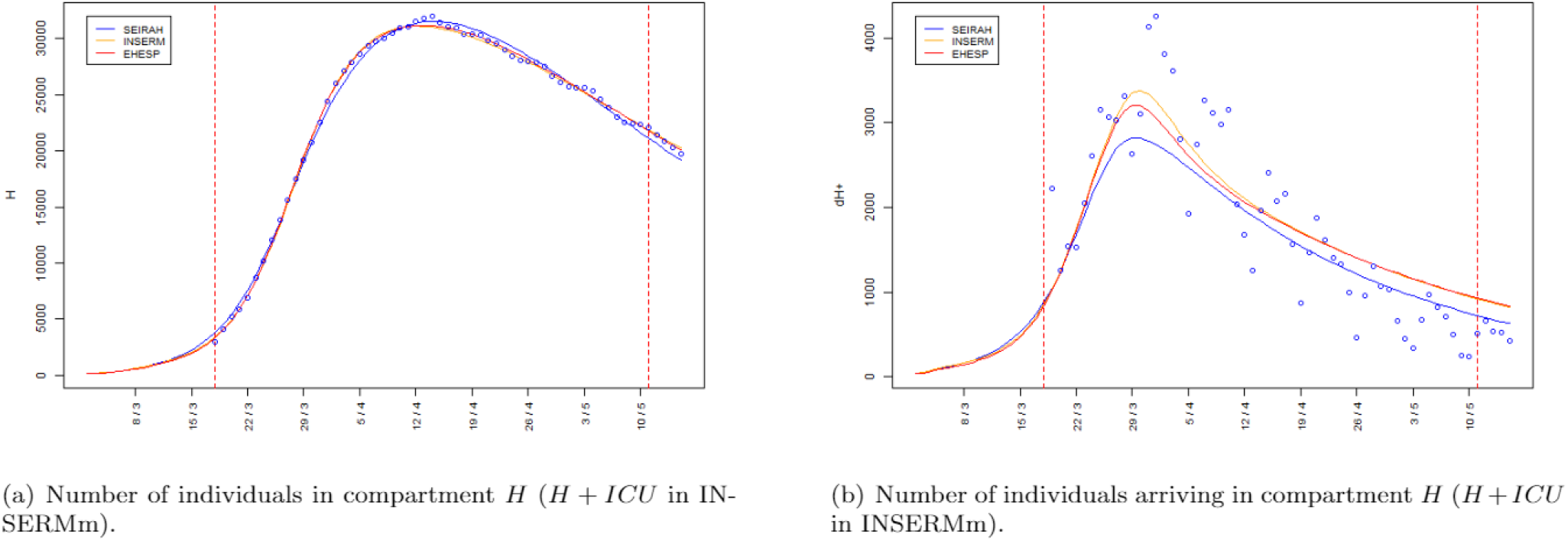
Hospitalization data over time in whole France as observed (blue circles) and computed by each model.

The three models provided similar results regarding the daily prevalent numbers of individuals in compartment H, but not regarding compartments *A, I, E*, or *R* (see the curves for the compartments *I, E*, and *R* in Figure B.1, Appendix B). Despite an apparent equivalence, there were much less individuals in compartment *I* with SEIRAHm than in compartments *I* + *I*_*nh*_ with EHESPm or *I*_*ps*_ + *I*_*ms*_ + *I*_*ss*_ compartments with INSERMm (Figure 3). Similarly, the cumulative number of removed individuals in compartment *R* (*R* + *D* in INSERMm) was much higher with INSERMm and EHESPm than with SEIRAHm. The numbers in compartment *D*, as predicted by INSERMm, were accurate vs. the official numbers published by the French government; however, the predicted prevalence for compartment *ICU* was almost twice higher than the observed one (Figure B.2 in Appendix B).

**Figure 3:**
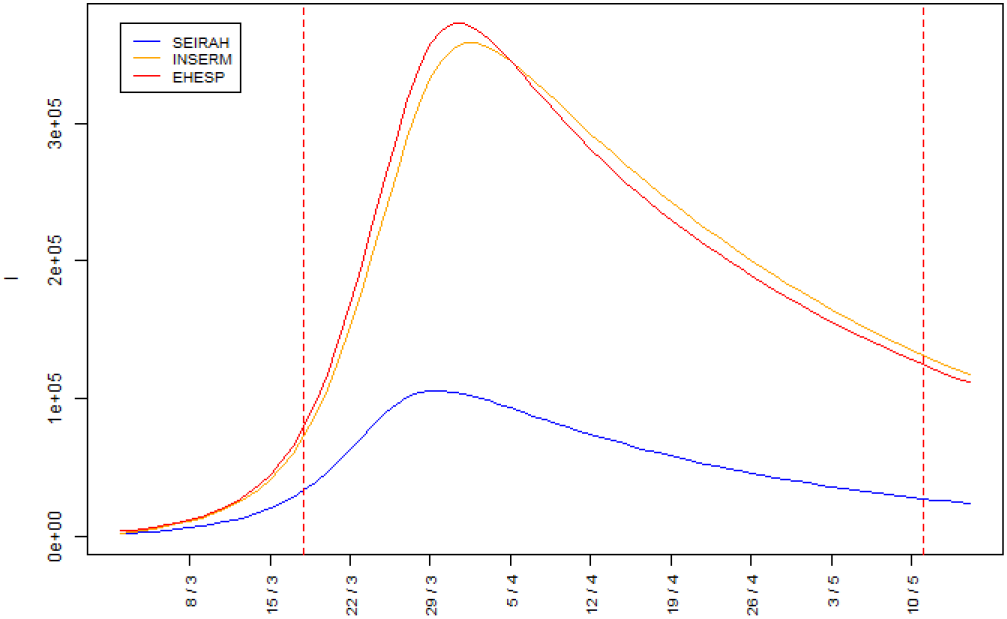
Number of individuals in compartment *I* over time in whole France (*I* in SEIRAHm, *I* + *I*_*nh*_ in EHESPm and *I*_*ps*_ + *I*_*ms*_ + *I*_*ss*_ in INSERMm).

The results obtained for compartments *H, E, I*, and *R* at a regional scale were similar to those obtained at the national scale (Figure B.1, Appendix B). Estimated parameter *β*_1_ was always lower but parameter *β*_2_ was always higher with SEIRAHm vs. EHESPm or INSERMm. Estimated parameter *E*_0_ with SEIRAHm was often very different from the one estimated with the two other models, but it could be higher (regions ARA and SUD) or lower (regions IDF and GE). With the exception of GE (and, to a lesser extent, IDF), parameters *T*_*c*_ were close with the three models. All parameters estimated for the INSERMm and EHESPm were generally similar.

### 3.2 Prediction of hospitalizations during the lockdown period

When the three models fitted the numbers of hospitalized patients as provided by SPF at the start of the lockdown (March 18 to April 6, 2020, F-SL approach), the national predictions for period April 7 to May 11 showed a strong overestimation of hospitalization prevalence with INSERMm and EHESPm but a satisfactory estimation with SEIRAHm (Figure 4). The overestimation was associated with a delay in the peak of the prevalence of hospitalized cases. The estimated number of individuals in compartment *E*_0_ at start of modelling was also three times lower with SEIRAHm than with the two other models (Table B.2). However, parameter *β*_1_ was much higher with SEIRAHm than with the two others and barely higher with EHESPm than with INSERMm. Parameter 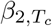 was smaller with SEIRAHm than with the two other models; this could have contributed to the smaller number of predicted hospitalized cases at the end of lockdown by this model. The values of *T*_*c*_ estimated by the three models were 24 to 25 days.

**Figure 4:**
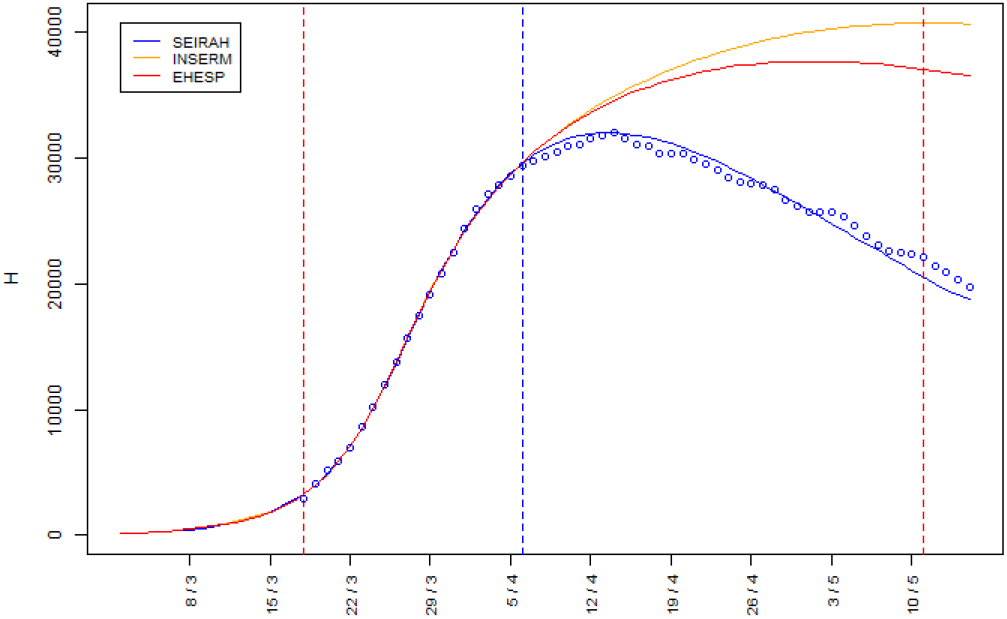
Number of individuals in compartment *H* over time in whole France as computed by each model (*H* + *ICU* in INSERMm).

The predictions of regional indicators were quite heterogeneous. All models overestimated the daily numbers of hospitalized (prevalent) cases in IDF. For ARA, just like EHESPm, INSERMm performed well but SEIRAHm overestimated the maximum daily number of hospitalized cases then underestimated the daily number of hospitalized cases at the end of the lockdown. For GE, SEIRAHm overestimated and EHESPm underestimated slightly the prevalence of hospitalized cases, whereas INSERMm was rather accurate. For SUD, EHESPm predicted satisfactorily the number of daily hospitalized cases, whereas SEIRAHm underestimated it and INSERMm overestimated it. Except for ARA, the estimated number of prevalent hospitalized cases at the beginning of the modelling was lower with SEIRAHm than with the two other models (Table B.2), whereas parameters *β*_1_ were higher with SEIRAHm than with the two other models and barely higher with EHESPm than with INSERMm. Parameters 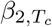 were smaller with SEIRAHm than with the two other ones; this may explain the smaller number of predicted hospitalized cases at the end of lockdown for with SEIRAHm, except for the IDF region. GE excluded, the values of *T*_*c*_ estimated by the three models for the three other regions were quite close. Each overestimation of the number of prevalent hospitalized cases was generally associated with a delayed corresponding peak and the underestimation with an advanced peak. In all regions, the maximum number of individuals in hospital was observed after April 6, whereas the parameters were estimated on purpose before the peak of hospitalizations was reached.

### 3.3 Predicted epidemic course after lockdown

When the F-WL approach was applied to fit the national data, the *R*_0_ estimated with INSERMm and EHESPm were very close (0.8 and 0.9 respectively; Table 1), whereas the one estimated with SEIRAHm was higher (= 1.3). Figure 5 shows that the scenario retained by SEIRAHm overestimated the number of hospitalized individuals after June 10. The scenarios obtained with SEIRAHm underestimated the hospitalization prevalence after the lockdown during the second half of May (Figure B.4a), which was not the case with INSERMm or EHESPm (Figures B.4c and B.4e).

**Table 1:** Estimated *R*_0_

|  | France | ARA | IDF | GE | SUD |
| --- | --- | --- | --- | --- | --- |
| SEIRAHm F-WL | 1.4 | 0.7 | 1.5 | 1 | 1.5 |
| INSERMm F-WL | 0.8 | 0.1 | 1.2 | 0.8 | 0.9 |
| EHESPM F-WL | 0.9 | 0.2 | 1.4 | 0.9 | 1 |
| SEIRAHm F-ALEL | 0.9 | 0.2 | 1.2 | 1.3 | 1.3 |
| INSERMm F-ALEL | 0.8 | 0.3 | 1.2 | 0.7 | 1.1 |
| EHESPM F-ALEL | 0.9 | 0.4 | 1.3 | 0.7 | 1.1 |
| Government estimations<sup>a</sup> | 0.93 | 1.02 | 0.91 | 0.89 | - |

Estimated $R_0$ with F-WL and F-ALEL approaches on June 6, 2020.
<sup>a</sup>The weekly estimations made by the French government and published for the week between 06/06/2020 and the 12/06/2020. Due to the fact that "SUD" is the aggregate of 3 regions, there is no associated government estimation.

**Figure 5:**
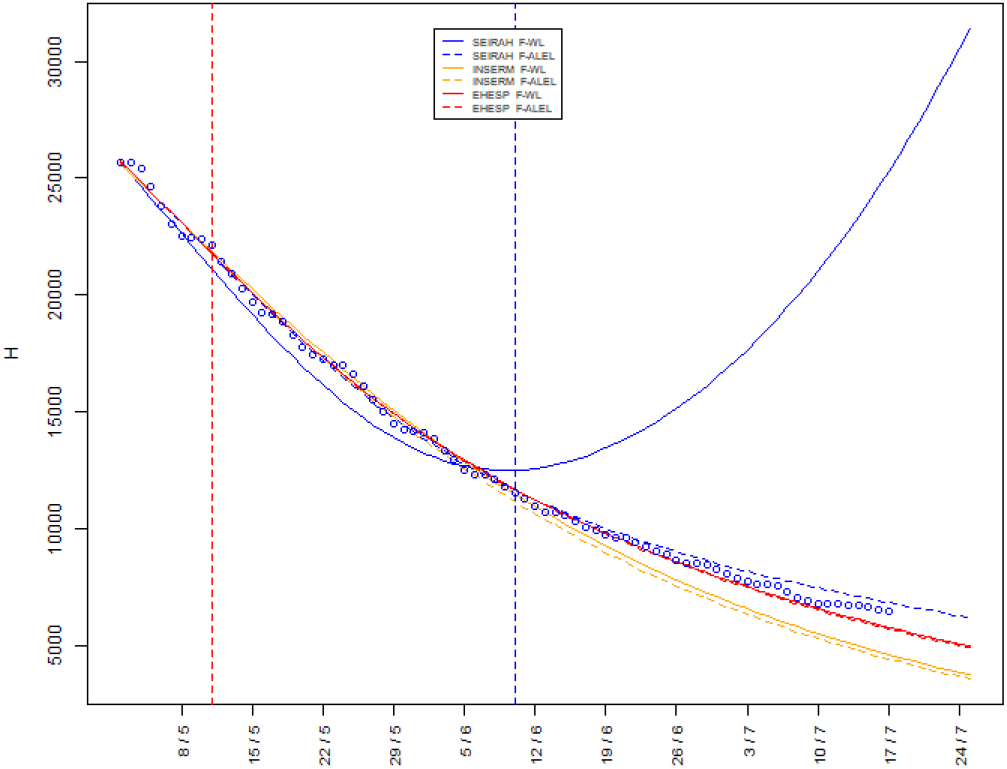
Number of individuals in compartment *H* over time in whole France as computed by each model (*H* + *ICU* in INSERMm). Results obtained with F-WL approach are represented by solid lines and results with F-ALEL approach by dotted lines.

When the F-WL approach was applied to fit the regional data, SEIRAHm estimated a higher *R*_0_ than INSERMm and EHESPm (Table 1). The *R*_0_ estimated by fitting EHESPm were slightly higher than those estimated with INSERMm. The hospitalization prevalence after June 10 indicated that the *R*_0_ estimated by SEIRAHm was clearly overestimated for regions IDF, GE, and SUD but slightly underestimated for ARA. The predictions of INSERMm and EHESPm were quite reasonable for regions GE and SUD, overestimated for IDF, and underestimated for ARA (Figure B.5, Appendix B.2).

When the F-ALEL approach was applied to fit the national data, SEIRAHm showed a much better fit than with the F-WL approach (Figure 5, dotted lines). The three models estimated the same *R*_0_ for June 2 (= 0.9). The *R*_0_ estimated with this approach during the lockdown was 0.66 with SEIRAHm, 0.77 with INSERMm, and 0.8 with EHESPm (Table B.4). Figure B.4b shows that SEIRAHm scenarios were more realistic than those obtained with F-WL approach. On the contrary, INSERMm and the EHESPm provided similar estimations with F-WL and F-ALEL approaches (Figures B.4d and B.4f). Regarding the results obtained with the F-ALEL approach during the early stage of the epidemic, all three models predicted too many hospitalizations at the beginning of the lockdown, which was not the case at the end of the lockdown (Figure B.6).

When the F-ALEL approach was applied to fit the regional data, the three models led to close results for SUD region, but quite different for GE region. For all regions except GE, the three models estimated a higher E0 and a lower *β*_1_ with F-ALEL approach than with F-WL approach (Table B.3).

## 4 Discussion

### 4.1 Description of the epidemic during the whole lockdown period

With F-WL approach, all three models succeeded in matching the observed prevalence in compartment *H*. However, regarding the estimated parameters (Table B.1, Appendix B.1), several comments may be made: (i) The estimated values of *T*_*c*_ for whole France, (24.3 to 26.2 days after the start of the simulation on March 2) corresponds to a delay of 8.3 to 10.2 days from the beginning of the lockdown, which is reasonable given the incubation period of the disease and the time needed to establish the lockdown; (ii) The equations of the three models correspond to specific functional forms and lead to different parameter estimates. The high value of *β*_1_ obtained with SEIRAHm allowed explaining the differences between estimated values of the parameters by the three models. Indeed, a high *β*_1_ means a high virulence that requires few infected individuals at time *t* = 0 (low *E*_0_) to fit hospitalization data. In such a case, the daily incidence rate needs to be strongly shrunk to reflect the lockdown effect (low 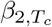 and short *T*_*c*_). Regarding the regions (GE with SEIRAHm excluded), the values of *T*_*c*_ ranged from 22.7 to 27.9 and were thus quite similar to those of whole France. Point (ii) remains true for the four regions.

Whereas the models were built to match the daily prevalence of hospitalized cases, the estimated prevalence in other compartments differed by a factor of 2 to 4 at both national and regional levels (Figure B.1, in Appendix B.2). In particular, the number of symptomatic cases (compartment *I* in SEIRAHm, *I*_*ps*_ + *I*_*ms*_ + *I*_*ss*_ in INSERMm, and *I*+*I*_*nh*_ in EHESPm) was more than three times lower with SEIRAHm than with the two other models. This admits two explanations. The first one is that, unlike INSERMm and EHESPm, SEIRAHm did not consider age groups; thus, the mean stay in compartment *H* of SEIRAHm was approximated as a weighted mean of age-specific mean stay times in compartment *H* from the two other models, the weights being the age-specific probabilities of hospitalization (see Equation (13) in Appendix A.2). However, with this simplification, it was not possible to obtain age-independent models able to reproduce the dynamics of age-dependent models. Indeed, the proposed simplification ignores possible changes of average age in compartment *H* over time (due to the fact that the proportion of individuals of each age group in compartment *H* change over time), resulting in a lack of flexibility. These parameter differences explain also the lower estimation of daily hospital incidences in Figure 2b. The second explanation is the structural differences between the models. The incidence of hospitalized cases (in compartment *H*) depends directly on the prevalence of symptomatic cases (in compartment *I*). The incidence in compartment *H* was calculated as the positive term of compartment *H* derivative which in SEIRAHm is given by System 4:

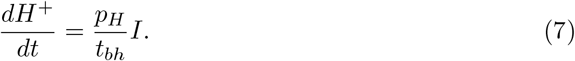

In INSERMm, the same incidence is given by System 5 (summing incidence in *H* and *ICU*):

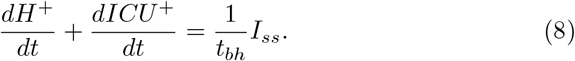

Finally, in EHESPm, the incidence given by System 6 is:

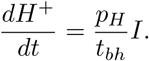

Despite the differences linked to the fixed parameters explained in Section 2.2.1 (Paragraph INSERM model), the incidence of hospitalized cases was nearly the same with the three models (Figure 2b). Thus, the numbers *p*_*H*_*I* with SEIRAHm, *I*_*ss*_ with INSERMm, and *p*_*H*_*I* with EHESPm were rather close. However, in SEIRAHm, compartment *I* was the only one that represented the infected symptomatic cases, whereas INSERMm included *I*_*ms*_ and *I*_*ps*_, and EHESPm compartment *I*_*nh*_. Excluding the parametrization effect related to the average age in compartment *H*, comparing the number of infected symptomatic cases in SEIRAHm and EHESPm is direct: in EHESPm, this number is higher by an amount *I*_*nh*_ than in SEIRAHm (this interpretation is further investigated in C.2). The comparison with INSERMm is not straightforward making it necessary to rewrite the equations for *I*_*ss*_ and *I*_*ms*_ + *I*_*ps*_ as follows:

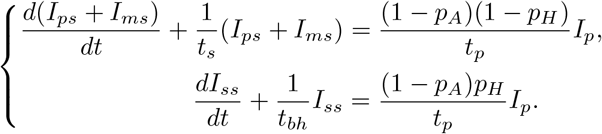

Assuming that the flows are much larger than the population variation^[6]^ in the left-hand side of both equations, the system can be rewritten:

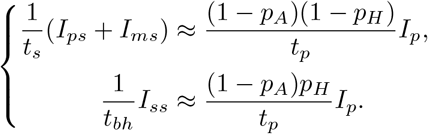

Combining the two equations leads to:

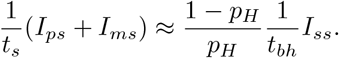

As the incidences of hospitalized cases predicted by the three models are close (see Figure 2b), using Equations (7) and (8) leads to:

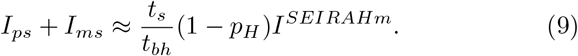

The coefficient 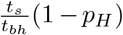 is roughly equal to 2, which explains much of the observed difference^[7]^. The differences in observed infected symptomatic cases implies differences between the infectious populations of the models.

This contributes to the higher *β*_1_ found with SEIRAHm while fitting this model on data of hospitalization. Indeed, given the lower number of infectious individuals, a higher virulence is needed to match the observed number of hospitalizations (see Equation 1 that reflects the entrance in the disease process where the second sum is the whole infectious population).

Each compartment may be subject to structural differences. As shown in Appendix C, it is not possible to make an intuitive equivalence between compartments from two different models due to the non-stationary regimes of the three models. Thus, obtaining mathematically and medically consistent parameters that allow the dynamics of a model to be reproduced by a model with a different structure is almost impossible. The impossibility to replicate the results of one model by another raises a tricky issue: how can one be sure that the chosen model is the most suitable in a given context? This is a crucial question because structural differences come with a huge difference in the number of recovered individuals (Figure B.1 in Appendix B), and may thus lead to take very different measures to contain the epidemic. Disappointingly, it is impossible to tell which model was the best during the first stage of the COVID-19 epidemic because of the lack of suitable data. Thus, any model designed to predict the recurrence of an epidemic should be written with extreme care and caution and all hypotheses made on the disease courses should be validated against observed population-based data.

This study results can be compared with results available in the literature. In particular, *R*_*e*_ (computed as de-scribed in Appendix A.4) and the prevalence of *R* over time were plotted in Figure B.7 along with various published results for France. The values obtained here are within the ranges reported by the literature though the latter vary widely. This heterogeneity makes it impossible to prefer one model to another.

The three models used here were able to match hospital data but disagreed regarding almost all other compartments. The structure of each model, from age classes to compartment layouts, led to very different predictions regarding the course of the epidemic. These differences may have major effects on prediction: in exploring the risk of recurrence or computing *R*_*e*_, the number of already immune individuals has a great impact on health policy.

Testing regularly a representative sample of individual allows monitoring the real number of infectious individuals (PCR tests) or the real number of recovered individuals (serological tests) over time and ensures more efficient model calibration. This is strongly recommended at various scales (local, regional, national); well-sized stratified samples would really help understanding the dynamics of the epidemic at each level.

### 4.2 Prediction of the course of the epidemic during the lockdown period

The French national predictions of prevalent hospitalized cases during the lockdown were strongly overestimated by INSERMm and EHESPm, whereas SEIRAHm predictions were close to the observed data (Figure 4). With the F-WL approach as with the F-SL approach, parameter *β*_1_ was almost two times higher with SEIRAHm than with the two other models, whereas the 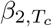 with SEIRAHm was smaller.

Regarding the regional models (Figure B.3 in Appendix B.2), only the predictions with INSERMm for ARA and GE, and those of EHESPm for SUD were accurate. Overall, the quality of the results was unstable; it varied greatly between models and regions. All models indicated an important effect of lockdown (with all models, 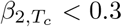, see Table B.2 in Appendix B.1),but failed to reproduce the F-WL estimations of the parameters. The average relative difference in estimated *β*_1_ was 5.1% between F-WL(Table B.1) and F-SL(Table B.2) and of more than 20% for parameter 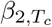. The sole difference between F-WL and F-SL was the length of the time interval used to fit the models; it showed that it was not sufficient to fit compartment *H* over the 20 first days of the lockdown. Moreover, the differences were larger re-garding 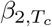 that needs a longer duration to seize the effect of the lockdown. Indeed, the full effect of 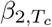 on the incidence in compartment *E* is obtained 24.5 days^[8]^ after March 2 (*t* = 0), which corresponded to March 26; and, as it takes on average 8.1 days (i.e., *t*_*i*_ + *t*_*bh*_) to reach *H* from *E*, this means that the full effect of 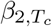 on the incidence in *H* was likely to be noticed after April 3. The fact that, with F-SL approach, the models were fitted using data from March 18 to April 6 may explain the large difference in estimated 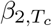 vs. F-WL approach.

The use of such compartmental models does not seem appropriate to predict the course of the epidemic during a lockdown when fitted only on data on compartment *H* at the beginning of the lockdown. Again, this underlines the need for prevalence data on compartment *I* that can lead to a better estimation of the lockdown effect at an early stage of the epidemic.

### 4.3 Predicted epidemic course after exiting the lockdown

In this part of the study, the model parameters have been first fitted before the end of the lockdown (on the daily prevalence of hospitalizations from March 18 to May 11 with F-WL approach and from March 8 to March 18 plus from May 1 to May 11 with F-ALEL approach). The models have then been used for projections with different values of post-lockdown *R*_0_ (Figure B.4) over a short term (May 11 to June 10) and medium term (June 11 to July 25). Next, with each model, the *R*_0_ maximizing the profile likelihood over the short term was retained.

The results concerning the post-lockdown scenarios obtained with F-WL were mixed. However, at a national level, the post-lockdown *R*_0_ values of 0.8 and 0.9 (Table 1, corresponding respectively to post-lockdown *R*_*e*_ values of 0.73 and 0.82) obtained with EHESPm and INSERMm were in line with the value 0.84 of the *R*_*e*_ given by the French government on its official dashboard on June 4 (obtained using the daily number of individuals visiting the emergency units as per the SurSaUD database). They were also in good agreement with other literature results (Table B.4) such as [10] (*R*_0_ value of 0.99 on May 30 and 0.89 on June 8 obtained with a simple SEIR model and fitting the SPF daily numbers of confirmed incident cases). These estimations are close despite methodological differences at two main levels: (i) the data used were different; and, (ii) the models used different representations of the disease process. With SEIRAHm, the post-lockdown *R*_0_ value obtained was much higher than with EHESPm or INSERMm, probably because F-WL approach was fitted using all lockdown data. This use of all data provided more information on the overall lock-down but led to an underestimation of the number of hospitalizations at lockdown end (Figure B.4a).This underestimated number of prevalent hospitalized cases led, in the short term, to the selection of a scenario that corresponded to an overestimated *R*_0_ (Figure 5).

At the regional level, all three models with F-WLL approach provided some unrealistic estimates of *R*_0_ (Table 1); either too low (ARA) or too high (IDF). As for SEIRAHm at the national level, this error could be due to the fitting method that did not favor a perfect match with end-of-lockdown data. It could also be due to the models that assumed linear variations for *β*_2,*t*_ at end of lockdown, which could be unrepresentative of the true variations of *R*_0_.

F-ALEL approach improved significantly the results with SEIRAHm at the national scale (Figure 5), but not much those obtained with INSERMm and EHESPm. The overestimation of the prevalent hospitalized cases (vs. SPF values) observed at the beginning of the lockdown on Figure B.6 may be attributed to the use of SurSaUD database before the lockdown for parameter estimation. Indeed, the correction of SurSaUD data (for better coherence with SPF data) may have been insufficient because of a non-constant corrective coefficient linked with the non-uniform distribution of SurSaUD database hospitals over the French territory.

At the regional level over a short term (May 11 to June 10), almost all prevalent hospitalizations as estimated with the F-ALEL approach were closer to the observed values than those estimated with the F-WL approach (Figure B.5). However, F-ALEL approach did not systematically improve the estimates of *R*_0_. Indeed, the models performed better regarding hospitalizations at end of lockdown but *R*_0_ variations could not be more accurately captured.

Thus, at both regional and national levels, F-ALEL seemed more appropriate than F-WL to build profiles for short-term predictions but both failed to provide medium-term predictions. Here too, knowing the prevalence in other compartments (such as *I* or *R*) could improve the quality of model calibration despite the need for a more complex approach to account for *R*_0_ variations after a lockdown in medium-term predictions.

## 5 Conclusion

Compartmental models have been used worldwide since the beginning of the COVID-19 pandemic to estimate or predict various parameters linked to its course in the population with the hope of helping decision-making regarding hospital management and/or lockdown and exit strategies. The designs, analyses, and results of these models are useful for all epidemiologists to adapt, extend, refine, or interpret their own models.

The analysis of these three models highlighted the influence of model structure on predictions. Whereas the three models described correctly the daily number of hospitalized cases and allowed estimating the changes in parameter values during the COVID-19 pandemic with data on only 20 days, they had limited abilities in predicting changes of the number of hospitalized cases during and after the lockdown period. As the information was restricted to hospitalized cases, intensive care cases, and disease related deaths, the information on the epidemic provided by the study of these data alone did allow knowing precisely the extent of the epidemic. As the information was restricted to hospitalized cases, intensive care cases, and disease related deaths, the information on the epidemic provided by the study of these data alone did allow knowing precisely the extent of the epidemic.

These compartmental models were designed to fit epidemic data rather than predict the course of the epidemic. Regular tests on representative samples of the population would make it possible to monitor precisely the number of infected individuals (PCR tests) and immune individuals (serological tests) and better seize the dynamics of the epidemic. The seroprevalence of COVID-19 infection in May-June 2020 analyzed by an INSERM study [11] provided important information on the dynamics of the epidemic before and during the lockdown and on the risk factors associated with past COVID-19 infections. Repeating this analysis over time should provide further details on more recent dynamics of the epidemic, whereas a complementary study of the dynamics of PCR positive tests is crucial to understand the more recent course of the epidemic and predict soon enough the occurrence of a potential new wave.

The analysis of the predictive properties of compartmental models would greatly benefit from comparisons of the results obtained after adjustments with results from simulations with multi-agent models.

## Data Availability

All data used during this study are available on data.gouv.fr or in the published articles listed in the references, except APHP related data obtained from a personal communication of the EHESP. All data generated during this study are available from the corresponding author on reasonable request.

http://data.gouv.fr

## 6 Abbreviations

COVID-19: Coronavirus disease 2019
ICU: Intensive care unit
SEIRAHm: Susceptible-Exposed-Infectious-Recovered-Asymptomatic-Hospitalized model
NSERMm: INSERM model of the Institut National de la Santé et de la Recherche Médicale
EHESPm: model of École des Hautes Études en Santé Publique
F-WL: Fit over the whole lockdown
F-SL: Fit at start of lockdown
F-ALEL: Fit ante lockdown and end of lockdown
ARA: Auvergne-Rhône-Alpes region
GE: Grand Est region
IDF: Île-de-France region
SUD: Nouvelle Aquitaine, Occitanie, and Provence-Alpes-Côte d’Azur regions
SPF: Santé publique France
SurSaUD database: “Surveillance sanitaire des urgences et des décès” database
APHP: Assistance publique – Hôpitaux de Paris.

## 7 Acknowledgements

The authors thank the research teams from the EHESP, INSERM and Université de Bordeaux for their cooperation and advise. The authors also thank the EHESP for sharing their codes and data.

## 8 Authors’ contribution

NP and RG implemented the three models and ran the simulations. RG, NP, CP, CR, VV, SD, JPB, AB, POG, AEL, SP, SL, PR participated to the study design, the statistical analysis and wrote the manuscript. SL and PR coordinated the project. JI contributed to editing the manuscript. All authors read and approved the final version of the manuscript.

## 9 Authors’ information

Not applicable

## 11 Ethics approval and consent to participate

Not applicable.

## 12 Consent for publication

Not applicable

## Appendix A: Supplementary materials

This appendix presents additional figures, the values of the parameters, the calibration process, and the method used for computing *R*_0_ and *R*_*e*_.

### A.1 Compartment initialization

Epidemiological models require often the calibration of a starting parameter. This parameter may be either the date of the beginning of the epidemic or the number of people in some compartments. Originally, EHESPm was calibrated using the first method, whereas SEIRAHm and INSERMm were calibrated using the second method. For a fair comparison between models, we calibrated the three models using the second method. Thus, all three models were supposed to begin on March 2, 2020^[9]^. In the following, this date corresponds to *t* = 0 (days).

To minimize the number of calibrated parameters and lessen the chance of overfitting, only one compartment was calibrated: the population in compartment *E* on March 2. The other compartments were initialized as follows. Let *Z*(*t*) denotes the number of individuals at time t in a compartment *Z*, and Δ*H*^+^(*t*) denotes the number of cases entering compartment *H* during day *t* (entering aggregation of *H* and *ICU* in INSERMm). Only SurSaUD database had data at the end of February / beginning of March. It contains only the value of Δ*H*^+^(*t*) over time, and as in [5], the hypothesis that nobody has exited from the *H* compartment before March 2 was made. The first hospitalization recorded by SurSaUD database occurred on February 27, 2020; thus, this hypothesis seems reasonable. Whence, the number of hospital beds occupied at the end of day 0 is 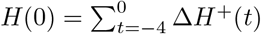.

It was also assumed that, in the early period of the pandemic, the number of dead or recovered individuals was negligible vs. the overall population and thus *R*(0) was set to 0 (and also *D*(0) in INSERMm).

Furthermore, there is a high volatility in the SurSaUD database, which was even truer at the beginning of the epidemic. Consequently, the data were smoothed using a moving average^[10]^.

The complete formulas that compute the initial population in the compartments of the three models are given in Table A.1. To give the intuition behind these formulas, let us consider a sub-part of EHESPm (see Figure A.1).

**Figure A.1:**
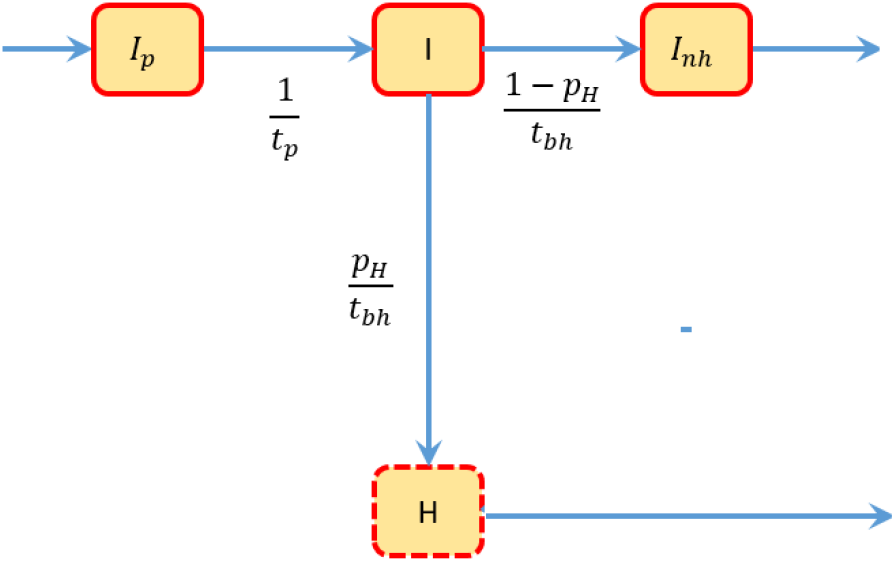
Sub-part of EHESPm.

The dynamics of compartments *I, H* and *I*_*nh*_ are given by

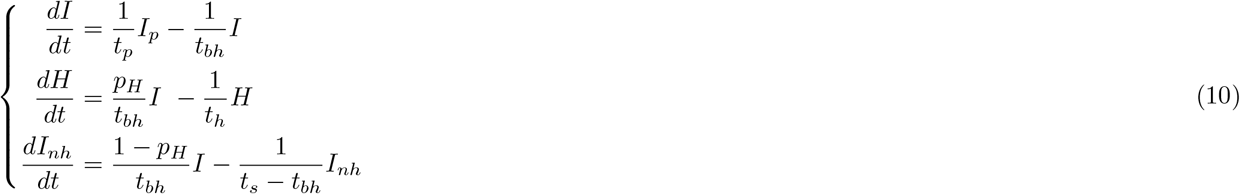

Since *p*_*H*_, *t*_*bh*_, *I, H* and *t*_*h*_ are non-negative, using the second equation of system 10, the number of cases entering compartment *H* during day *t* can be approximated by 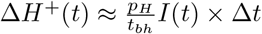, with Δ*t* = 1 day. Thus

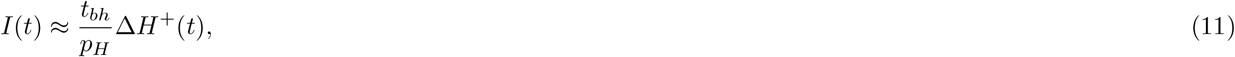

and *I*(0) is then set to 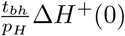.

The first equation of system 10 can be rewritten as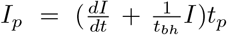. Then, the approximation 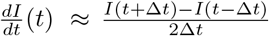 leads to 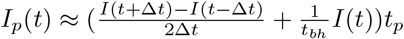.

For *t* = 0 and Δ*t* = 1, using equation 11, *I*p(0) is then set to 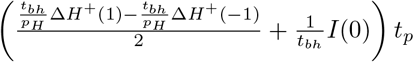.

Since *I*_*nh*_ and (*t*_*s*_*− t*_*bh*_) are non-negative, using the third equation of system 10, the number of cases entering compartment *I*_*nh*_ during day *t* can be approximated by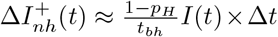, with Δ*t* = 1 day. Then, equation 11 leads to

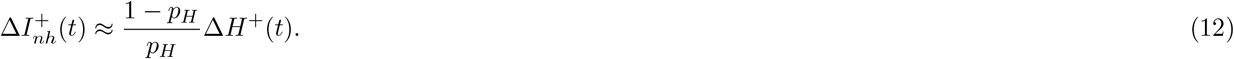

Supposing that nobody has exited from compartment *I*_*nh*_ before March 2, as for compartment *H*, then 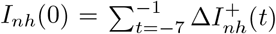. Thus, by equation 12, 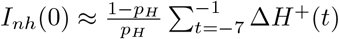. So, *I*_*nh*_(0) is set to 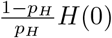.

The other compartments are derived in a similar way. For models stratified by age, *H*(0) and other compartments are first computed as an aggregation for all age groups. Then, their partitioning is made according to the distribution of age in the number of hospital beds occupied on March 18 as reported by Santé Publique France.

### A.2 Parameters

Most of the parameters were chosen according to the literature. The parameters directly linked to the contagiousness of the disease over time (i.e. *β*_1_, *β*_2,*t*_ and *T*_*c*_) were estimated (see Section A.3). Parameter *E*(0) was also estimated, while other compartment initial values were derived from the SurSaUD database (see Section A.1). Limiting the number of estimated parameters reduces the risk of overfitting, and having the same number of parameters for each model allows a fair comparison between them. A summary of the parameters used can be found in Table A.2. The values of age-dependent parameters can be found in Table A.3. For consistency, when a parameter was detailed by age group in INSERMm and EHESPm, a weighted mean (using the population of each age group as weight) was calculated to provide the corresponding parameter value in SEIRAHm.

**Table A.1:** Initial populations in the compartments.

| Parameters | SEIRAHm | INSERMm | EHESPm |
| --- | --- | --- | --- |
| Starting date (t=0) | 02-03-2020 | 02-03-2020 | 02-03-2020 |
| $S(0)$ | $N_{tot} - \sum_{Z=E,A,I\dots} Z(0)$ | $N_{tot} - \sum_{Z=E,A,I\dots} Z(0)$ | $N_{tot} - \sum_{Z=E,A,I\dots} Z(0)$ |
| $E(0)$ | Estimated | Estimated | Estimated |
| $I_p(0)$ | - | $\left( \frac{t_{bh}\Delta H^+(1) - t_{bh}\Delta H^+(-1)}{2} + \frac{I_{ss}(0)}{t_{bh}} \right) \frac{t_p}{(1-p_A)p_H}$ | $\left( \frac{\frac{t_{bh}}{p_H}\Delta H^+(1) - \frac{t_{bh}}{p_H}\Delta H^+(-1)}{2} + \frac{I(0)}{t_{bh}} \right) t_p$ |
| $A(0)$ | $(I(0) + H(0)) \frac{p_A}{1-p_A}$ | $I_{ss}(0) \frac{p_A}{(1-p_A)p_H}$ | $(I_p(0) + I(0) + I_{nh}(0) + H(0)) \frac{p_A}{1-p_A}$ |
| $I(0)$ | $\Delta H^+(0) \frac{t_{bh}}{p_H}$ | - | $\Delta H^+(0) \frac{t_{bh}}{p_H}$ |
| $I_{ps}(0)$ | - | $I_{ss}(0) \frac{p_{Ips}}{p_H}$ | - |
| $I_{ms}(0)$ | - | $I_{ss}(0) \frac{p_{Ims}}{p_H}$ | - |
| $I_{ss}(0)$ | - | $\Delta H^+(0) t_{bh}$ | - |
| $I_{nh}(0)$ | - | - | $H(0) \frac{1-p_H}{p_H}$ |
| $H(0)$ | $\sum_{t=-4}^0 \Delta H^+(t)$ | $(1 - p_{ICU}) \sum_{t=-4}^0 \Delta H^+(t)$ | $\sum_{t=-4}^0 \Delta H^+(t)$ |
| $ICU(0)$ | - | $p_{ICU} \sum_{t=-4}^0 \Delta H^+(t)$ | - |
| $D(0)$ | - | 0 | - |
| $R(0)$ | 0 | 0 | 0 |

**Table A.2:** Parameter valuesTable

| Parameters | Description | SEIRAHm | INSERMm | EHESpm | Source |
| --- | --- | --- | --- | --- | --- |
| $\beta_1$ | Daily incidence hazard rate given a contact with someone contagious | | Estimated | | - |
| $T_c$ | Date where the lockdown is fully effective | | Estimated | | - |
| $\beta_2, T_c$ | Change of $\beta_1$ during lockdown | | Estimated | | - |
| $t_i$ | Total incubation time | | 5.1 days | | [5], [7], [12], [13] |
| $p_A$ | Probability of being asymptomatic | | 0.5 | | [6], [14] |
| $i_Z$ | Relative infectivity. Set 0.5 for asymptomatic, to 0 for compartments S, E, H, ICU, R and D. Set to 1 for all other infectious compartments. | | $i_A = 0.5$ | | [6], [15] |
| $t_s$ | Time before recovering (compartments A and I) | | 7 days | | [16]<sup>a</sup> |
| $t_{bh}$ | Time before hospitalization | | 3 days | | [2], [17] |
| $p_H$ | Probability of hospitalization | 0.08 | By age | | EHESP<sup>c</sup>, [7] |
| $t_h$ | Hospitalization duration | 17.3 | By age | | EHESP<sup>c</sup> |
| $N_{tot}$ | Total population | By region | By age and region | | [7], INSEE |
| $C$ | Contact matrix | - | By age | | [7], [8] |
| $t_p$ | Duration of the prodromic phase | - | 1.5 day | | [6], [16]<sup>a</sup> |
| $p_{ICU}$ | Probability of admission to ICU while at hospital | - | By age | - | EHESP<sup>c</sup> |
| $t_{icu}$ | Time before recovering after admission to ICU | - | By age | - | EHESP<sup>c</sup> |
| $p_{D\vert H}$ | Probability of death at hospital (excluding ICU) | - | By age | - | EHESP<sup>c</sup> |
| $p_{D\vert ICU}$ | Probability of death while in ICU | - | By age | - | EHESP<sup>c</sup> |
| $p_{I_{ps}}$ | Probability of being paucisymptomatic | - | By age | - | Assumed<sup>b</sup> |
| $p_{I_{ms}}$ | Probability of developing moderate symptoms | - | By age | - | Assumed<sup>b</sup> |

<sup>a</sup> At the time this paper was written, there was no real consensus about this value in the specialized literature. A value coherent with reference [16] was then chosen.
<sup>b</sup> $p_{I_{ps}}$ and $p_{I_{ms}}$ were obtained supposing $p_{I_{ps}} = 1/3 \times p_{I_{ms}}$ under constraint $p_H + p_{I_{ps}} + p_{I_{ms}} = 1$ , using the values of $p_H$ given by age.
<sup>c</sup> Personal communication from the EHESP, whose team had access to APHP data, and provided the appropriate aggregated statistics.

The value of *p H* in SEIRAHm is given by 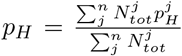, with *n* denoting the number of age groups, 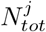 the population in age group *j*, and 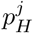 the probability of being hospitalized after showing symptoms for age group *j*.

The value of *t*_*h*_ in SEIRAHm is given by

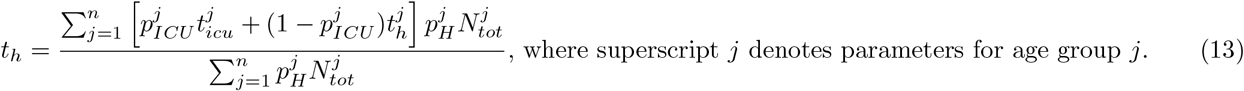

The 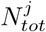 values were given by the French age distribution (source: INSEE). This may create a potential bias because some elderly people live in nursing homes and might not go to hospital. Examples of age distributions in the population are given in Table A.5 (for France and for ARA region).

**Table A.3:** Age-dependent parameters Values of the age-dependent parameters used in INSERMm and EHESPm, whenever applicable.

| age | 0-4 | 5-9 | 10-14 | 15-19 | 20-24 | 25-29 | 30-34 | 35-39 | 40-44 |
| --- | --- | --- | --- | --- | --- | --- | --- | --- | --- |
| $p_H$ | 0.001 | 0.001 | 0.003 | 0.003 | 0.012 | 0.012 | 0.032 | 0.032 | 0.049 |
| $p_{I_{ps}}$ | 0.25 | 0.25 | 0.249 | 0.249 | 0.247 | 0.247 | 0.242 | 0.242 | 0.238 |
| $p_{I_{ms}}$ | 0.749 | 0.749 | 0.748 | 0.748 | 0.741 | 0.741 | 0.726 | 0.726 | 0.713 |
| $p_{ICU}$ | 0.285 | 0.285 | 0.285 | 0.285 | 0.285 | 0.285 | 0.323 | 0.323 | 0.343 |
| $p_{D\vert ICU}$ | 0 | 0 | 0 | 0 | 0 | 0 | 0.2 | 0.2 | 0.31 |
| $p_{D\vert H}$ | 0 | 0 | 0 | 0 | 0 | 0 | 0.023 | 0.023 | 0.058 |
| $t_h$ | 8.1 | 8.1 | 8.1 | 8.1 | 8.1 | 8.1 | 7.4 | 7.4 | 8 |
| $t_{icu}$ | 19.8 | 19.8 | 19.8 | 19.8 | 19.8 | 19.8 | 20.7 | 20.7 | 20.5 |
| age | 45-49 | 50-54 | 55-59 | 60-64 | 65-69 | 70-74 | 75-79 | 80P |  |
| $p_H$ | 0.049 | 0.102 | 0.102 | 0.166 | 0.166 | 0.243 | 0.243 | 0.273 | |
| $p_{I_{ps}}$ | 0.238 | 0.225 | 0.225 | 0.209 | 0.209 | 0.189 | 0.189 | 0.182 | |
| $p_{I_{ms}}$ | 0.713 | 0.674 | 0.674 | 0.626 | 0.626 | 0.568 | 0.568 | 0.545 | |
| $p_{ICU}$ | 0.343 | 0.408 | 0.408 | 0.36 | 0.36 | 0.282 | 0.282 | 0.05 | |
| $p_{D\vert ICU}$ | 0.31 | 0.25 | 0.25 | 0.47 | 0.47 | 0.54 | 0.54 | 0.47 | |
| $p_{D\vert H}$ | 0.058 | 0.081 | 0.081 | 0.147 | 0.147 | 0.227 | 0.227 | 0.434 | |
| $t_h$ | 8 | 9.4 | 9.4 | 11.4 | 11.4 | 15.9 | 15.9 | 26.6 | |
| $t_{icu}$ | 20.5 | 24.5 | 24.5 | 17.6 | 17.6 | 23.4 | 23.4 | 19.2 | |

The contact matrix used in this study was computed as the sum of the five different contact matrices of [8] (matrices given for school, work, home, elderly, and others). It was also assumed that this contact matrix does not depend on the region under investigation. Considering those assumptions, the contact matrix can be summarized as in Table A.4. Notice that Prem et al. [8] considered 16 age groups, whereas this study, similarly with Roux et al. [7] considered 17 age groups, using in the contact matrix the same values for group 75-79 and group 80P (80 and older).

### A.3 Parameter estimation

Four parameters (i.e., *E*(0), *β*_1_, *β*_2,*t*_ and *T*_*c*_) have been estimated for each model by likelihood maximization. The prevalent hospitalized cases were supposed to follow a Poisson distribution. The models being different, several optimization methods were used and the best results selected for each model.

With SEIRAHm, a constrained Nelder-Mead method was applied using `constrOptim` function in R. With EHESPm and INSERMm, a first constrained Nelder-Mead fit was applied using package `lmfit` in Python. This method showed convergence difficulties for *E*(0) and *T*_*c*_. So, we retained the parameters obtained by a second fitting operation using package `emcee` in Python. This algorithm, detailed in [18], is an implementation of an affine-invariant ensemble sampler for Markov Chain Monte Carlo (MCMC).

**Table A.4:** Contact matrix used in this paper and issued from [8]. This representation is the transpose of contact matrix *C* in Equation 1.

|  |  |  |  |  |  |  |  |  |  |  |  |  |  |  |  |  |  |
| --- | --- | --- | --- | --- | --- | --- | --- | --- | --- | --- | --- | --- | --- | --- | --- | --- | --- |
| 80P | 0.05 | 0.05 | 0.07 | 0.03 | 0.05 | 0.02 | 0.05 | 0.05 | 0.05 | 0.08 | 0.09 | 0.09 | 0.13 | 0.14 | 0.33 | 0.64 | 0.64 |
| 75-79 | 0.05 | 0.05 | 0.07 | 0.03 | 0.05 | 0.02 | 0.05 | 0.05 | 0.05 | 0.08 | 0.09 | 0.09 | 0.13 | 0.14 | 0.33 | 0.64 | 0.64 |
| 70-74 | 0.09 | 0.06 | 0.07 | 0.03 | 0.05 | 0.03 | 0.05 | 0.10 | 0.09 | 0.08 | 0.09 | 0.12 | 0.26 | 0.28 | 0.97 | 0.37 | 0.37 |
| 65-69 | 0.13 | 0.11 | 0.09 | 0.05 | 0.05 | 0.05 | 0.09 | 0.15 | 0.11 | 0.09 | 0.12 | 0.22 | 0.43 | 1.00 | 0.62 | 0.37 | 0.37 |
| 60-64 | 0.19 | 0.18 | 0.10 | 0.07 | 0.08 | 0.11 | 0.18 | 0.26 | 0.18 | 0.15 | 0.27 | 0.51 | 1.41 | 0.52 | 0.66 | 0.28 | 0.28 |
| 55-59 | 0.27 | 0.19 | 0.15 | 0.16 | 0.33 | 0.36 | 0.41 | 0.36 | 0.27 | 0.34 | 0.65 | 1.46 | 0.68 | 0.44 | 0.34 | 0.34 | 0.34 |
| 50-54 | 0.31 | 0.21 | 0.27 | 0.48 | 0.81 | 0.96 | 0.81 | 0.92 | 1.11 | 1.09 | 1.92 | 0.92 | 0.44 | 0.32 | 0.34 | 0.54 | 0.54 |
| 45-49 | 0.31 | 0.33 | 0.57 | 1.05 | 1.27 | 1.00 | 1.14 | 1.34 | 1.68 | 2.20 | 1.74 | 0.68 | 0.42 | 0.30 | 0.43 | 0.44 | 0.44 |
| 40-44 | 0.50 | 0.85 | 1.08 | 1.02 | 0.97 | 1.26 | 1.46 | 2.10 | 2.95 | 1.57 | 1.59 | 0.79 | 0.52 | 0.43 | 0.51 | 0.45 | 0.45 |
| 35-39 | 1.04 | 1.14 | 0.83 | 0.76 | 1.13 | 1.47 | 1.89 | 3.21 | 1.88 | 1.47 | 1.21 | 0.75 | 0.69 | 0.45 | 0.48 | 0.38 | 0.38 |
| 30-34 | 1.21 | 0.92 | 0.40 | 0.53 | 1.17 | 1.78 | 2.92 | 1.76 | 1.59 | 1.26 | 1.18 | 0.92 | 0.64 | 0.45 | 0.25 | 0.31 | 0.31 |
| 25-29 | 0.75 | 0.42 | 0.24 | 0.65 | 1.71 | 3.46 | 1.72 | 1.41 | 1.25 | 0.99 | 1.29 | 0.93 | 0.51 | 0.31 | 0.24 | 0.19 | 0.19 |
| 20-24 | 0.34 | 0.16 | 0.29 | 1.36 | 3.93 | 1.94 | 1.01 | 0.76 | 0.92 | 0.95 | 1.02 | 0.61 | 0.34 | 0.21 | 0.16 | 0.15 | 0.15 |
| 15-19 | 0.26 | 0.27 | 0.85 | 7.83 | 2.17 | 0.73 | 0.42 | 0.68 | 1.11 | 1.54 | 1.20 | 0.75 | 0.36 | 0.16 | 0.36 | 0.36 | 0.36 |
| 10-14 | 0.41 | 1.01 | 6.99 | 2.30 | 0.26 | 0.14 | 0.53 | 0.84 | 0.98 | 0.61 | 0.83 | 0.57 | 0.26 | 0.26 | 0.31 | 0.45 | 0.45 |
| 5-9 | 1.11 | 5.03 | 1.54 | 0.31 | 0.17 | 0.25 | 0.85 | 1.07 | 0.66 | 0.47 | 0.59 | 0.67 | 0.36 | 0.31 | 0.29 | 0.32 | 0.32 |
| 0-4 | 3.80 | 1.06 | 0.24 | 0.12 | 0.20 | 0.53 | 0.72 | 0.70 | 0.31 | 0.41 | 0.25 | 0.54 | 0.40 | 0.20 | 0.11 | 0.24 | 0.24 |
|  | 0-4 | 5-9 | 10-14 | 15-19 | 20-24 | 25-29 | 30-34 | 35-39 | 40-44 | 45-49 | 50-54 | 55-59 | 60-64 | 65-69 | 70-74 | 75-79 | 80P |

**Table A.5:**
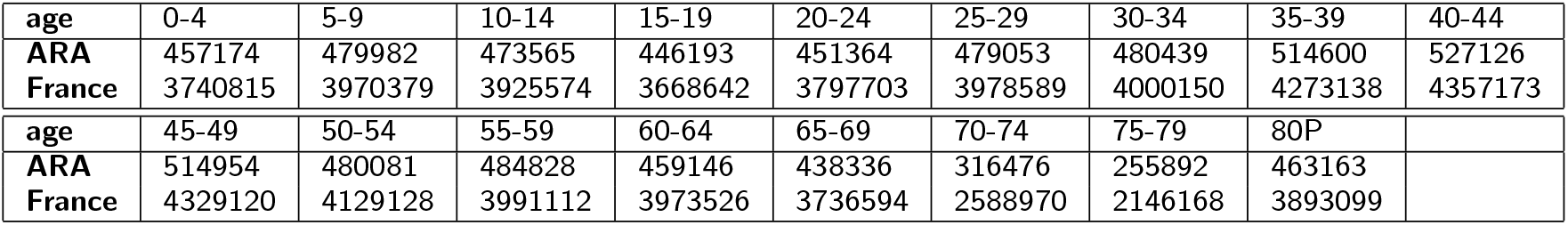
Examples of age distributions of the population.

In the three articles that introduced SEIRAHm, EHESPm, and INSERMm, the estimation of the parameters was always performed by likelihood maximization. However, the probability distributions and the data used for computing the likelihood were different. In this study, with all model, the likelihood was computed using the prevalent hospitalized cases, and assuming for them a Poisson distribution of a lambda parameter set at the number of hospitalizations of the day. In addition, for EHESPm, and INSERMm the *E*(0) compartment is assumed to be partitioned in age groups according to the age distribution of the population.

### A.4 *R*_0_ and *R*_*e*_

Two useful quantities in epidemiology are the basic reproductive number *R*_0_ and its associated effective reproduction number *R*_*e*_.

#### General concept

Quantity *R*_0_ can be seen as the average number of new infection cases caused by a single infected individual during her/his entire infectious period, in a completely susceptible population. To compute *R*_0_ as defined by Diekmann et al. [19], it is necessary to introduce the concept of next generation matrix. In the following, the approach and notational conventions of Van den Driessche and Watmough [20] are used. It is assumed that there is *m* different compartments of infected individuals, and *n* different age groups. Thus, there is a total of *nm* groups of infected (but not necessarily infectious) individuals. Let *x*(*t*) = (*x*_1_(*t*), *…, x*_*nm*_(*t*)), with quantity *x*_*i*_(*t*), 1 *≤ I ≤ nm*, denoting the number of individuals in the *i*th group of infected individuals at time *t*.

Let ℱ_*i*_(*x*(*t*)) be the rate of new infected individuals in the *i*th group of infected individuals at time *t*. Let 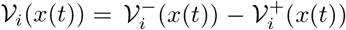 where 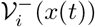 is the rate of departures from group *i*, and 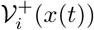 is the rate of arrivals in group *i* by all other means than new infections.

Let 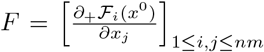 and 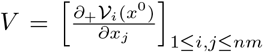 where *x*^0^ is a disease free equilibrium and *∂*_+_ denotes right partial derivatives. The next generation matrix is then defined as

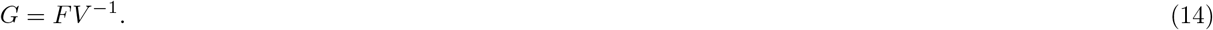

As shown in [20], 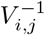 can be seen as the average time an individual in the *j*th group of infected individuals will spend in the *i*th group of infected individuals during his/her remaining lifetime. Thus, intuitively, the next generation matrix is the product of new individuals in each state over the average time an individual stays infectious. Finally, *R*_0_ is defined as the spectral radius of *G*.

The effective reproduction number *R*_*e*_ takes into account the number of remaining susceptible individuals. As defined by Brauer and Castillo-Chavez ([21], p.440), it is given by

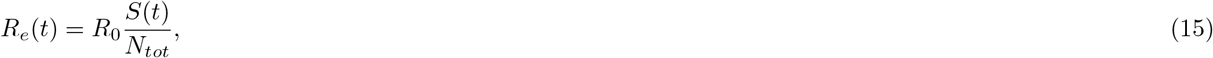

where *N*_*tot*_ is the total size of the population.

*R*_0_ *computation for SEIRAHm* This paragraph aims to illustrate the previous paragraph with the computation of the basic reproduction number *R*_0_ for SEIRAHm. In this model, there is no age group, and there are four compartments with infected individuals : *E, I, A, H*.

In the disease free equilibrium *x*^0^, we have *E* = 0, *I* = 0, *A* = 0, *H* = 0 and *S* = *N*_*tot*_, then the matrix *F* is given by

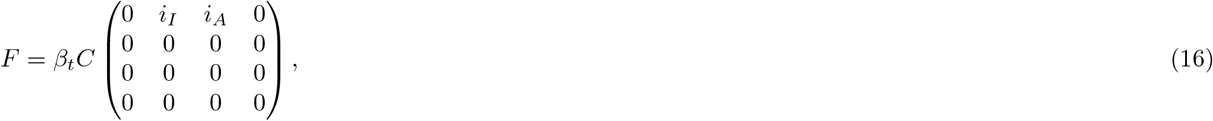

where *β*_*t*_ = *β*_1_*β*_2,*t*_ (cf. equations 1 and 2), and *C* is the mean number of daily contacts in the case of a single age group (cf. equation 2). The matrix *V* is given by

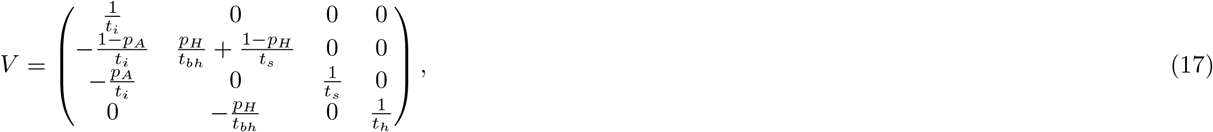

using Equation (14) and the definition of *R*, we have 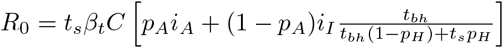.

#### Age group integration

Unlike SEIRAHm, both INSERMm and EHESPm have 17 different age groups. To define a common framework for both models, it was assumed that there are *m* different infectious compartments (for INSERMm, *m* = 8, and for EHESPm, *m* = 6), and *n* different age groups (*n* = 17 in both). The key hypothesis here is that movements across age groups are neglected: the total population 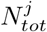 of age group *j* remains constant over time.

Let *x*(*t*) = (*x*_1,1_(*t*), *…, x*_*m,*1_(*t*), *x*_1,2_(*t*), *…, x*_*m,n*_(*t*)), with quantity *x*_*i,j*_(*t*) denoting the number of individuals in the *i*th compartment of infected individuals in the age group *j* at time *t*.

Let ℱ_*j*_(*x*(*t*)) = (ℱ_1,*j*_(*x*(*t*)), *…*, ℱ_*m,j*_(*x*(*t*))), with ℱ_*i,j*_(*x*(*t*)) denoting the rate at time *t* of new infected individuals belonging to age group *j* in compartment *i*. Matrix *F* can then be defined by sub-matrices *F*_*j*_1, _*j*2_, where *j*_1_ and *j*_2_ denote age group, that are the Jacobian matrices of ℱ_*j*_ with respect to variables 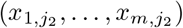 at *x*^0^.

Similarly, let *𝒱*_*j*_(*x*(*t*)) = (*𝒱*_1,*j*_(*x*(*t*)), *…, 𝒱*_*m,j*_(*x*(*t*))), with *𝒱*_*i,j*_(*x*(*t*)) denoting for age group *j* in compartment *i* the rate of departures minus arrivals for all other reasons than new infections. Matrix *V* is also defined by submatrices *V*_*j*_1 _, *j*_2 that are the Jacobian matrices of *𝒱*_*j*_ with respect to variables 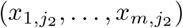 at *x*^0^.

*F and V for INSERMm.* Let 1 *≤ j*_1_, *j*_2_ *≤ n*. The expression of *F*_*j, j*_ ^[11]^ is given by

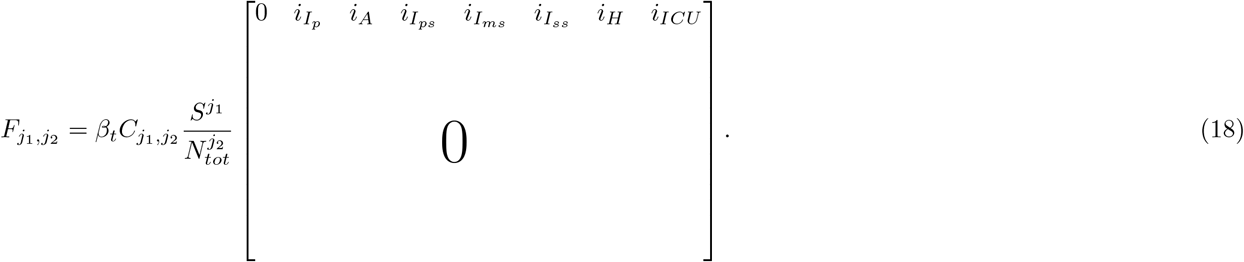

*V*_*j*1_, _*j*2_ is a null matrix if *j*_1_ */*= *j*_2_. When *j*_1_ = *j*_2_, let *j* = *j*_1_ = *j*_2_ then it is given by

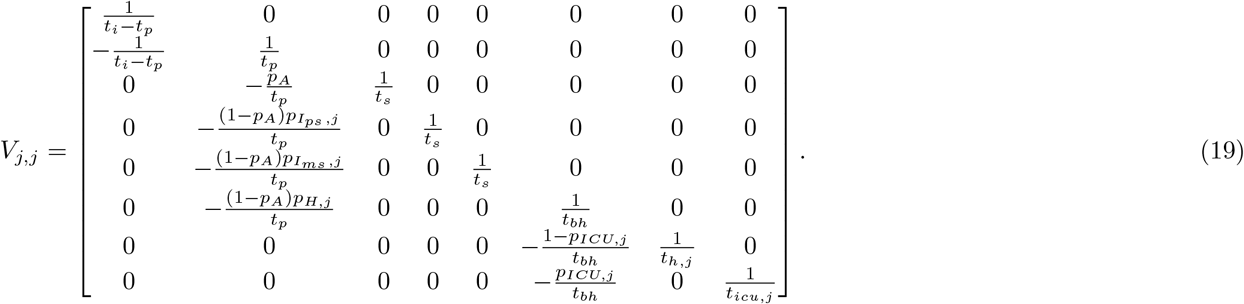

Here, *t*_*icu,j*_, *t*_*h,j*_, *pI*_*ms, j*_, *pI*_*ps, j*_, *p*_*H,j*_, *p*_*ICU,j*_ denote the values of the corresponding parameters for individuals in age group *j*. Again, *R*_0_ is given by the spectral radius of *FV*^−1^.

*F* and *V used to compute R0 for the EHESPm model* Let 1 *≤ j*_1_, *j*_2_ *≤ n*. The expression of *F j*_1_ = *j*_2_ ^[12]^ is given by

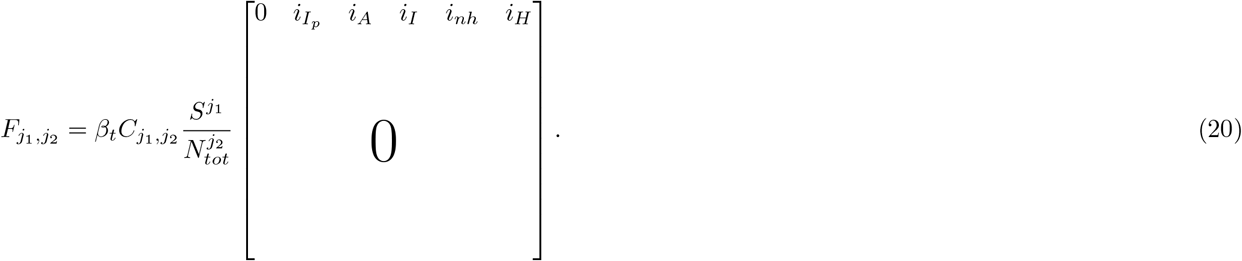

Again, *V*_*j*1_, _*j*2_ is a null matrix if *j*_1_ */*= *j*_2_. When *j*_1_ = *j*_2_, let *j* = *j*_1_ = *j*_2_, then *V*_*j,j*_ is given by

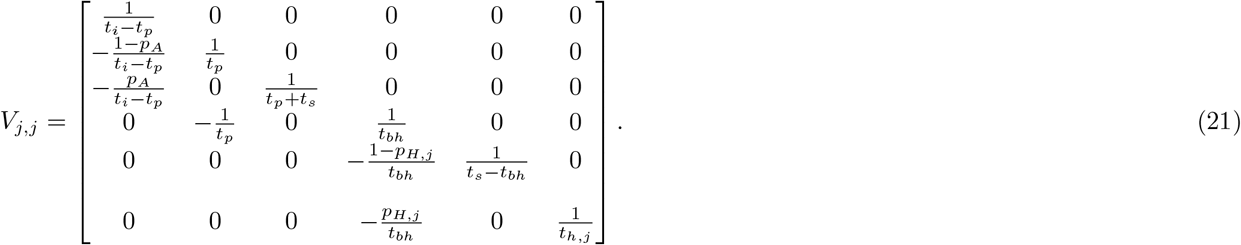

## Appendix B: Additional figures and results

### B.1 Estimated parameters

**Table B.1:** parameters as estimated with F-WL approach.

| | $E_0$ | | | $\beta_1$ | | | $\beta_{2, T_c}$ | | | $T_c$ | | |
| --- | --- | --- | --- | --- | --- | --- | --- | --- | --- | --- | --- | --- |
|  | SEI | INS | EHE | SEI | INS | EHE | SEI | INS | EHE | SEI | INS | EHE |
| France | 10 001 | 19 930 | 15 283 | 0.067 | 0.049 | 0.047 | 0.13 | 0.23 | 0.19 | 24.9 | 24.3 | 26.2 |
| ARA | 1 003 | 649 | 157 | 0.072 | 0.057 | 0.063 | 0.13 | 0.14 | 0.14 | 24.3 | 23.9 | 23.4 |
| IDF | 1 103 | 8 790 | 8 227 | 0.094 | 0.045 | 0.049 | 0.14 | 0.2 | 0.2 | 23.8 | 27.9 | 27.1 |
| GE | 1 014 | 8 506 | 8 138 | 0.104 | 0.039 | 0.043 | 0.14 | 0.22 | 0.22 | 16.9 | 23.5 | 22.7 |
| SUD | 997 | 248 | 37 | 0.068 | 0.049 | 0.05 | 0.11 | 0.15 | 0.16 | 26 | 26 | 26.1 |

**Table B.2:** parameters as estimated with F-SL approach.

| | $E_0$ | | | $\beta_1$ | | | $\beta_{2, T_c}$ | | | $T_c$ | | |
| --- | --- | --- | --- | --- | --- | --- | --- | --- | --- | --- | --- | --- |
|  | SEI | INS | EHE | SEI | INS | EHE | SEI | INS | EHE | SEI | INS | EHE |
| France | 4 955 | 15 345 | 12 252 | 0.076 | 0.046 | 0.049 | 0.13 | 0.25 | 0.23 | 25 | 24.3 | 24.3 |
| ARA | 996 | 756 | 337 | 0.071 | 0.056 | 0.06 | 0.1 | 0.14 | 0.13 | 26.6 | 24.4 | 24.4 |
| IDF | 1 115 | 6 921 | 6 725 | 0.095 | 0.048 | 0.053 | 0.24 | 0.28 | 0.26 | 21.6 | 24.5 | 24.3 |
| GE | 1 108 | 8 508 | 8 601 | 0.103 | 0.039 | 0.042 | 0.18 | 0.22 | 0.21 | 16.5 | 23.6 | 23.5 |
| SUD | 61 | 85 | 37 | 0.084 | 0.05 | 0.05 | 0.05 | 0.18 | 0.15 | 25.8 | 24.8 | 26.1 |

All estimated parameters of the three different models used in this paper are reported in Tables B.1, B.2 and B.3.

### B.2 Other figures

The results with F-WL approach for different compartments and all regions are given in Figure B.1. This figure shows the number of individuals in compartment *H* (*H* + *ICU* in INSERMm), compartment *I* (*I*+*I*_*nh*_ in EHESPm, *I*_*ss*_ + *I*_*ps*_ + *I*_*ms*_ in INSERMm), compartment *E* (*E* + *I*_*ps*_ in INSERMm and EHESPm) and compartment *R* (*R* + *D* in INSERMm). Compartment *A* is not printed, but the results were very close to those of *I*, except that EHESPm had more individuals in *A* than INSERMm because individuals could stay longer in *A* in EHESPm than in INSERMm.

**Table B.3:** Model parameters as estimated with F-ALEL approach.

| | $E_0$ | | | $\beta_1$ | | | $\beta_{2,T_c}$ | | | $T_c$ | | |
| --- | --- | --- | --- | --- | --- | --- | --- | --- | --- | --- | --- | --- |
|  | SEI | INS | EHE | SEI | INS | EHE | SEI | INS | EHE | SEI | INS | EHE |
| France | 15 328 | 29 719 | 28 435 | 0.068 | 0.043 | 0.046 | 0.15 | 0.19 | 0.2 | 20.7 | 24.5 | 23.7 |
| ARA | 1 018 | 1 641 | 1 380 | 0.069 | 0.047 | 0.05 | 0.13 | 0.16 | 0.17 | 25.2 | 27 | 26.3 |
| IDF | 4 539 | 12 021 | 12 309 | 0.077 | 0.043 | 0.047 | 0.15 | 0.21 | 0.21 | 21.8 | 26.8 | 25.7 |
| GE | 1 358 | 4 023 | 4 154 | 0.107 | 0.054 | 0.059 | 0.14 | 0.17 | 0.17 | 16 | 18.5 | 17.7 |
| SUD | 2 207 | 5 530 | 5 396 | 0.072 | 0.038 | 0.040 | 0.13 | 0.19 | 0.2 | 17.4 | 24.3 | 23.4 |

**Figure B.1:**
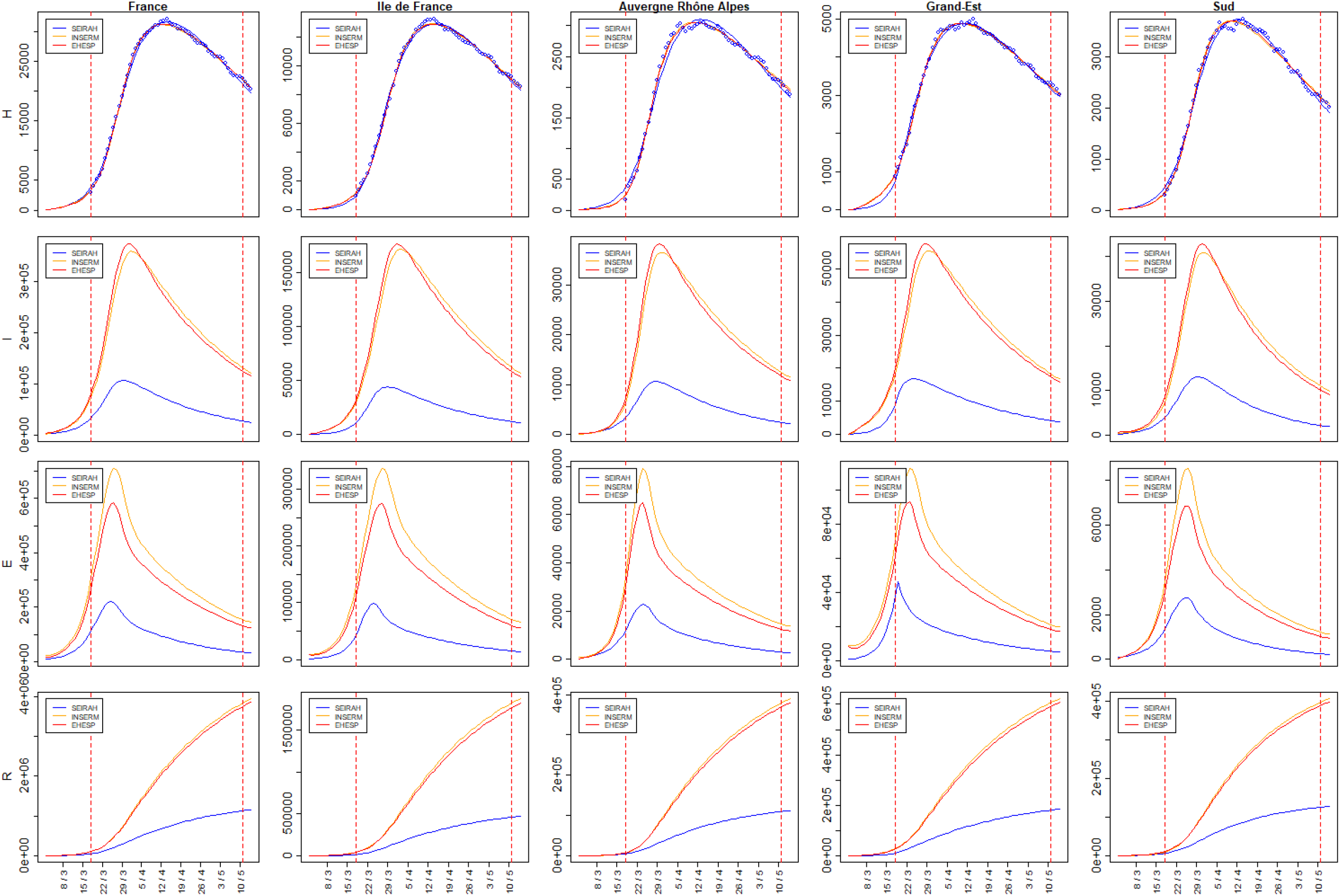
Number of individuals in aggregated compartments *H, I, E*, and *R* over time in whole France and the four regions as obtained with F-WL approach.

It was also possible to look at compartments *ICU* and *D*, which were specific to INSERMm. As shown in Figure B.2, the model seemed to grasp the number of individuals dead at the hospital over the fitting period, but it overestimated the number of individuals in compartment *ICU*. This could be expected because, in this model, compartment *ICU* includes also pre-ICU and post-ICU hospitalizations.

**Figure B.2:**
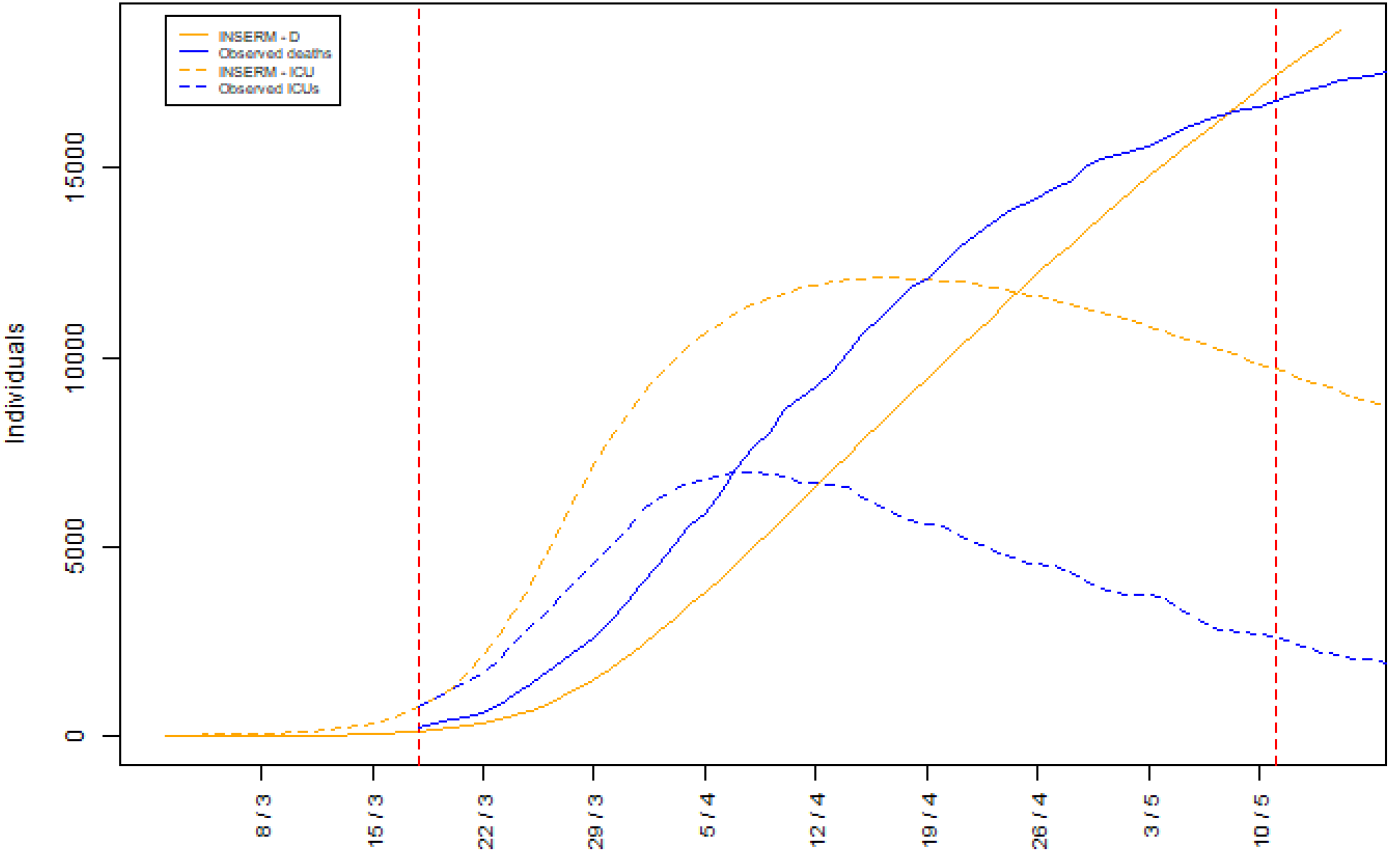
Number of individuals in compartments *ICU* and *D* in whole France. Observed values and values obtained with F-WL approach and INSERMm.

Figure B.3 shows the results of F-SL approach, for compartment *H* (*H* + *ICU* in INSERMm) in all regions. The scenarios obtained with the F-WL and the F-ALEL approaches with different *R*_0_ are given in Figure B.4 (compartment *H, H* +*ICU* in INSERMm for whole France) and the best scenarios for all regions are shown in Figure B.5. Figure B.6 presents the results for whole France with F-ALEL approach vs. SPF data (blue dots). In this approach, over the period before the lockdown, the model was fitted to corrected SurSaUD data. The difference between SPF data and the fitted model at the beginning shows that, even after correction, SurSaUD data hardly matched SPF data.

**Figure B.3:**
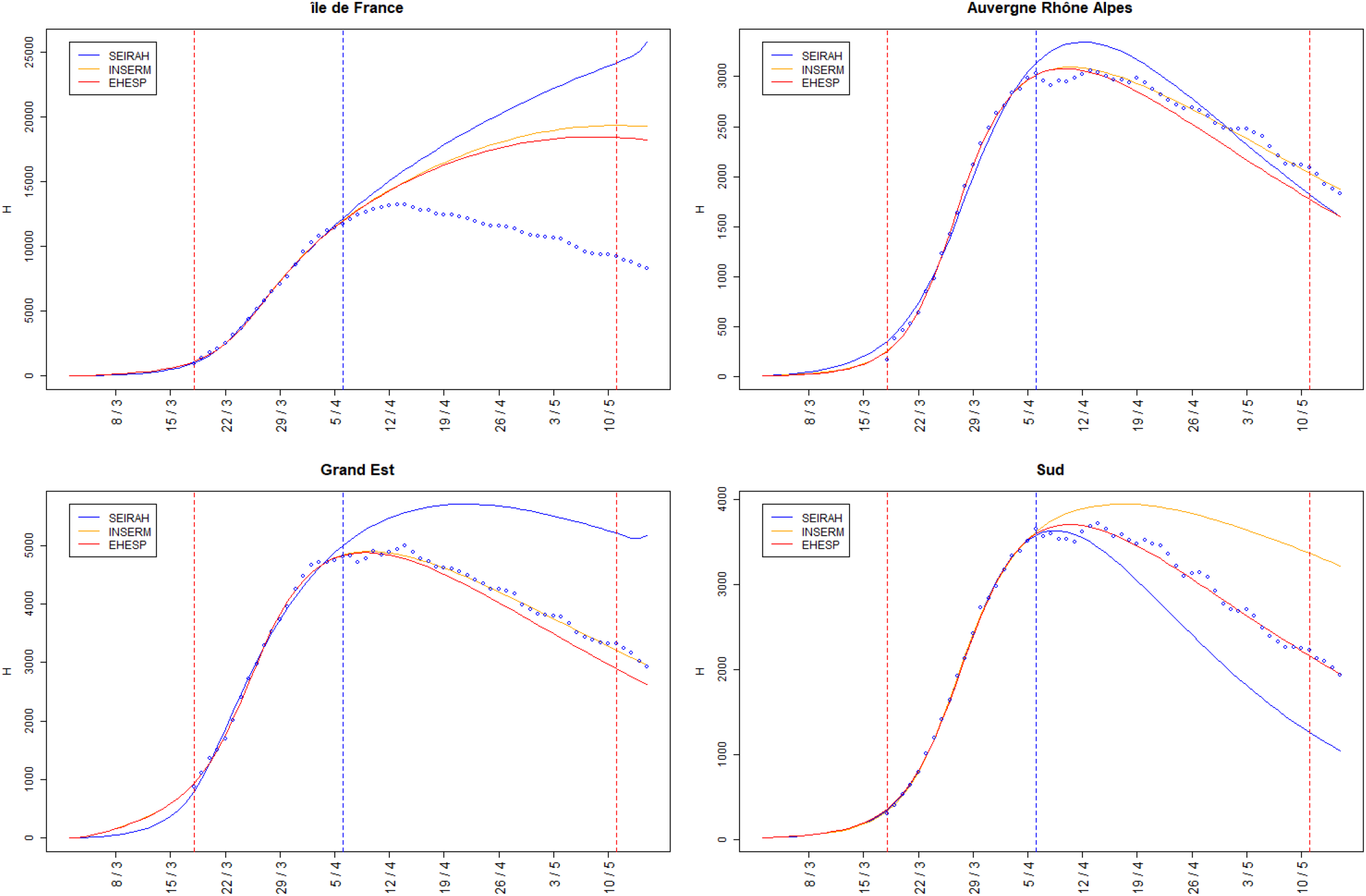
Number of individuals in compartment *H* (*H* + *ICU* in INSERMm) in the four regions, as obtained with F-SL approach.

**Figure B.4:**
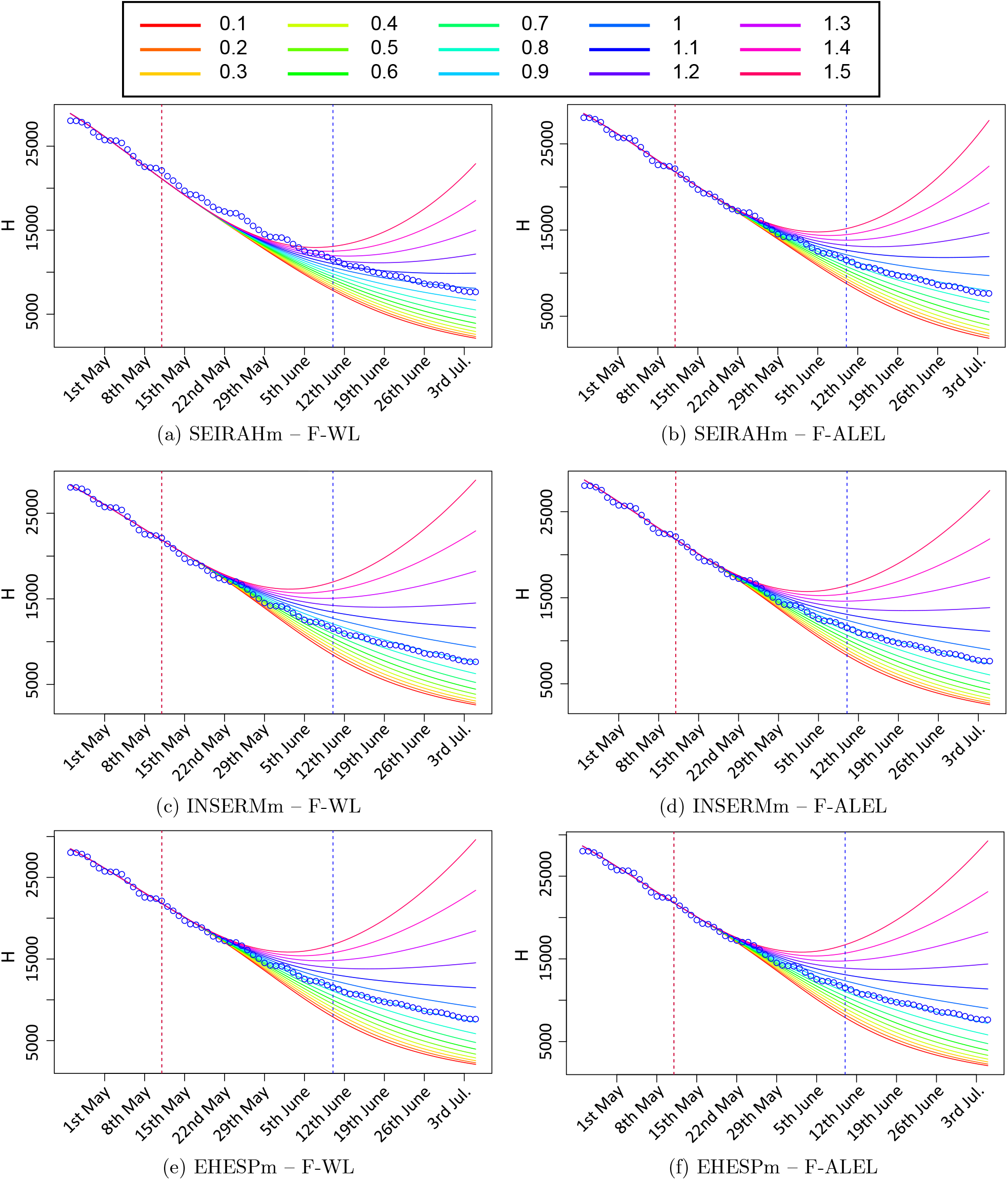
Number of individuals in compartment *H* (*H* + *ICU* in INSERMm) in whole France with different *R*_0_ scenarios as obtained with F-WL and F-ALEL approaches.

**Figure B.5:**
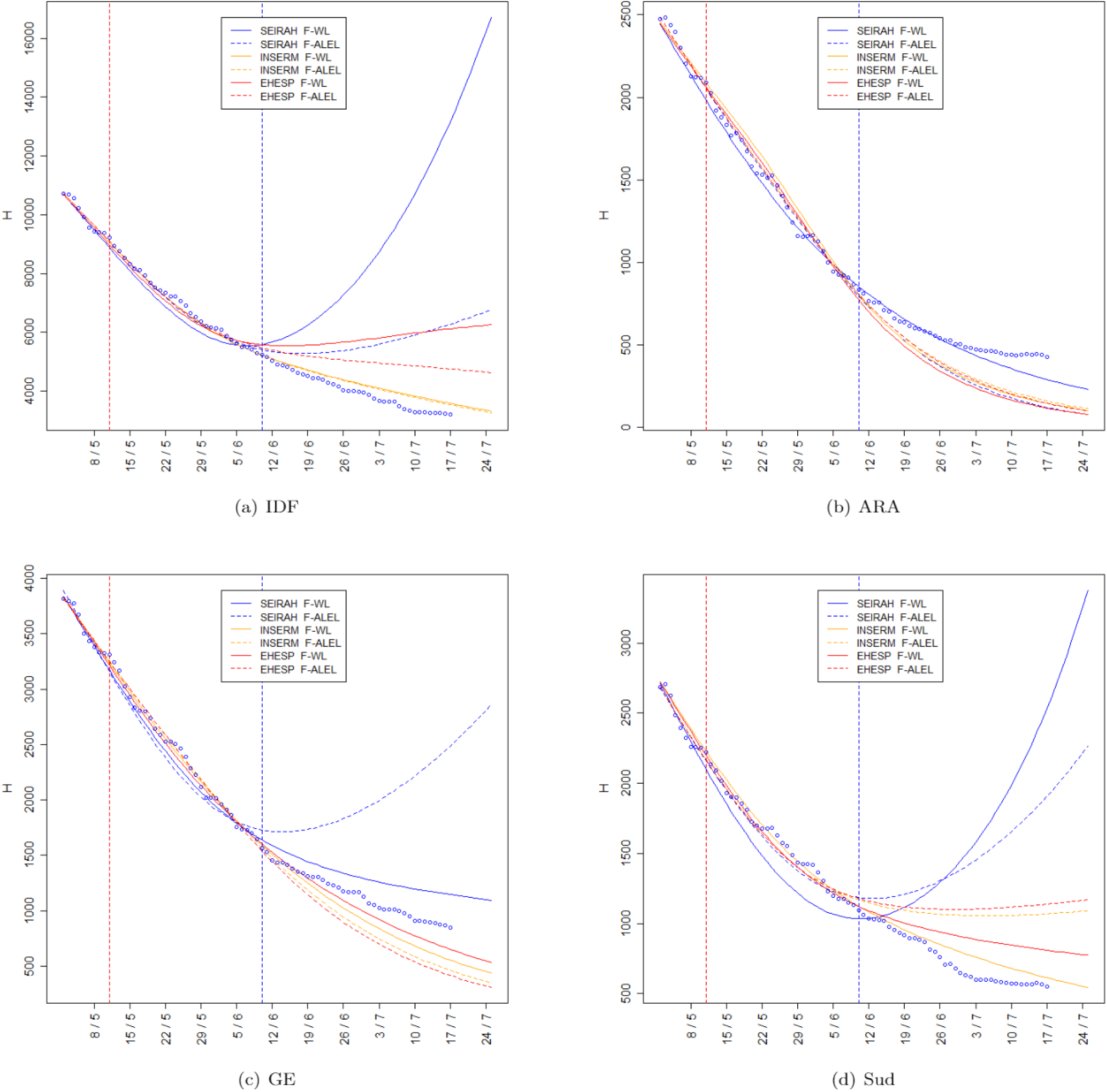
Number of individuals in compartment *H* (*H* + *ICU* in INSERMm) in each of the four regions with the best scenarios as obtained with F-WL and F-ALEL approaches.

**Figure B.6:**
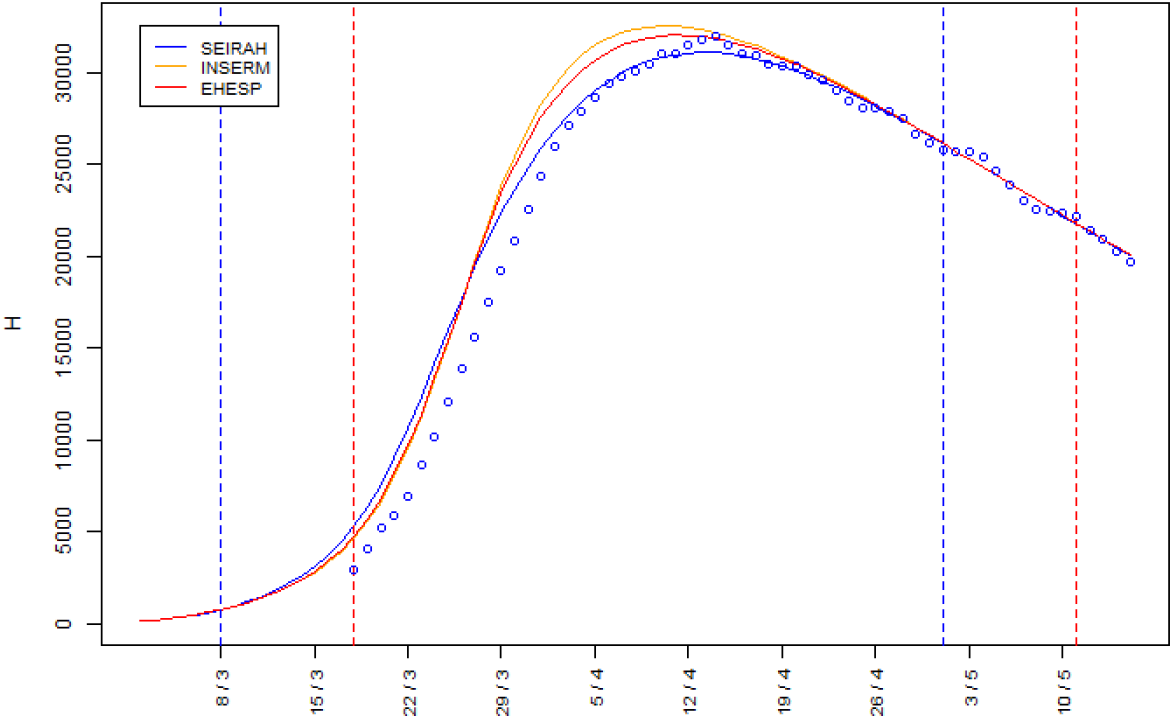
Number of individuals in compartment *H* over time (*H* + *ICU* in INSERMm) in whole France with each model as obtained with F-ALEL approach.

**Table B.4:**
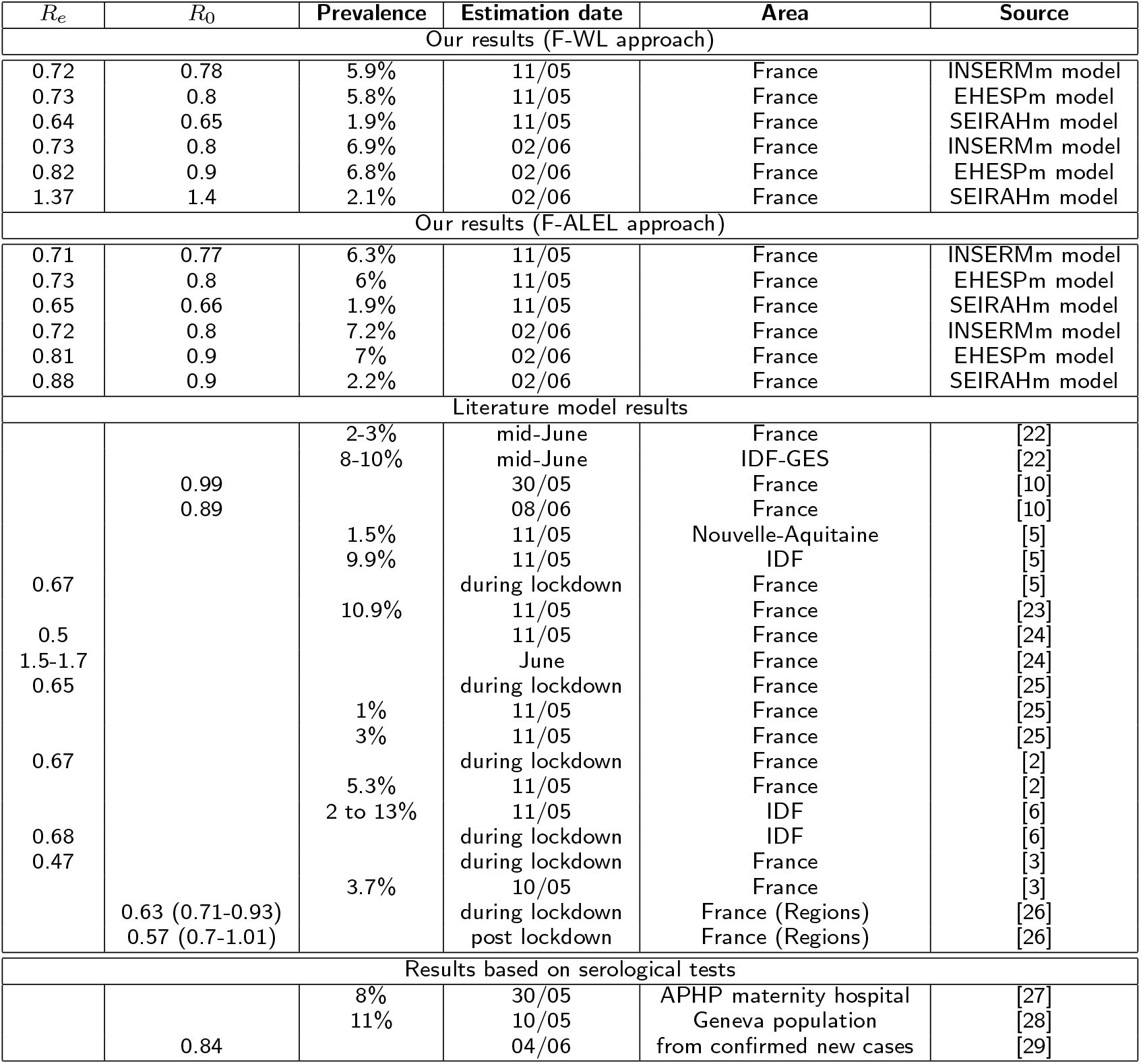
Present and literature results compared.

**Figure B.7:**
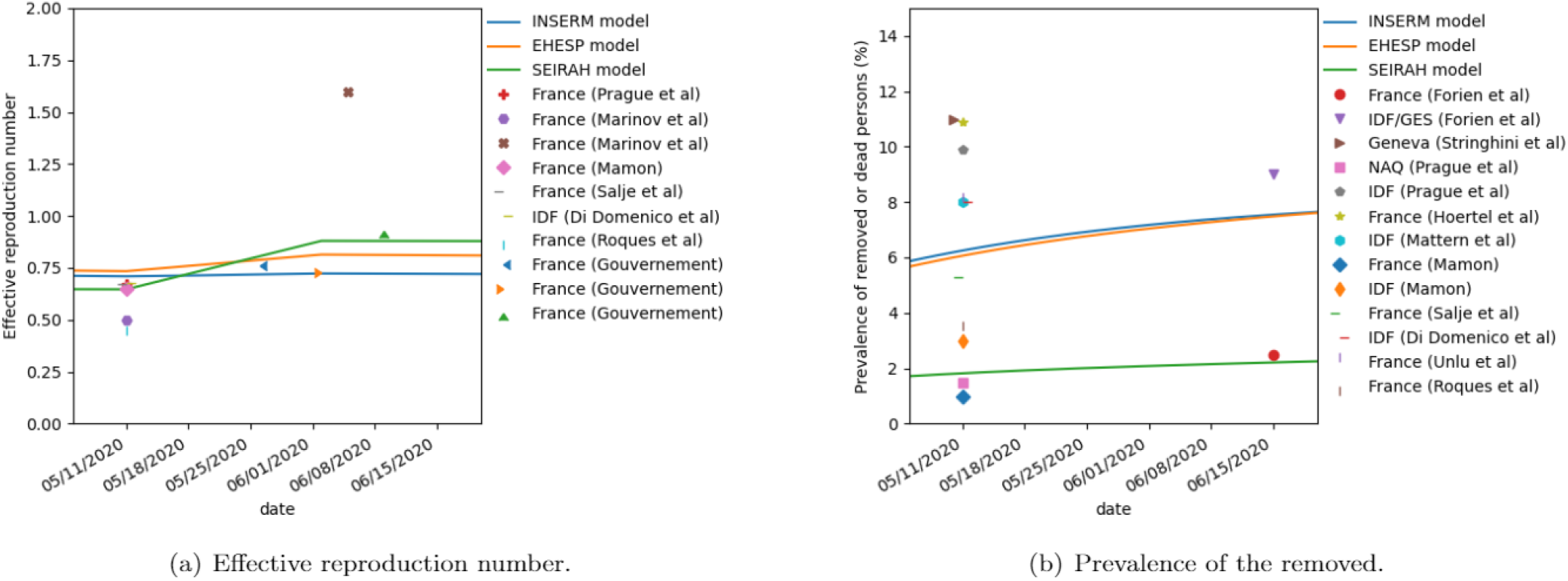
Comparisons between literature results and those of the present study (whole France, F-ALEL approach).

## Appendix C: On the importance of the architecture of compartmental models

### C.1 Toy models to understand differences between models

One big effort in this paper was the use of the same parameter set in three models. The probabilities and durations used to determine the transitions between compartments were combined to ensure consistency between models. For instance, the sum of the durations of the two compartments of the pre-symptomatic period in INSERMm and EHESPm was set to be equal to the duration of the pre-symptomatic compartment of SEIRAHm. However, the results of the different models still differed. To explain this, let us consider four compartmental toy models. The first and simplest one is:

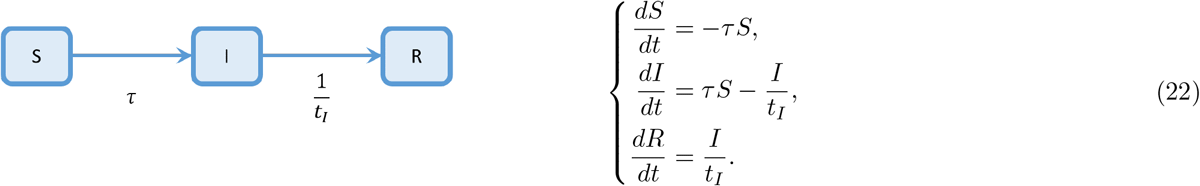

The following models can be seen as a detailed version of the first one. The second has two phases *I*_1_ and *I*_2_ during the infected period :

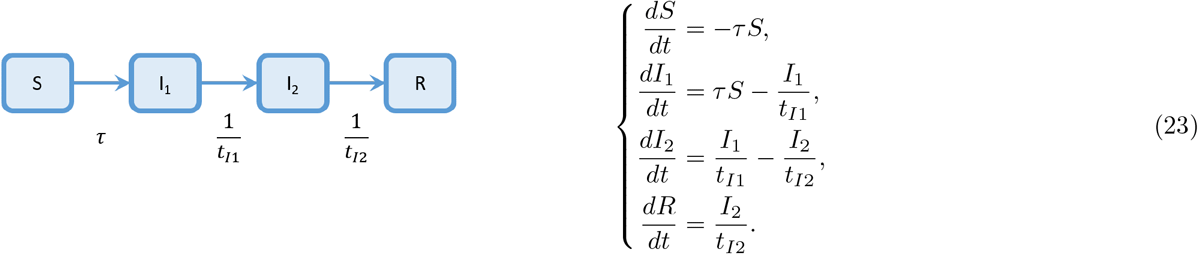

In the third model, there are two kinds of infected people: *I*_*A*_ and *I*_*B*_ with probabilities *p*_*A*_ and *p*_*B*_ (satisfying *p*_*A*_ + *p*_*B*_ = 1):

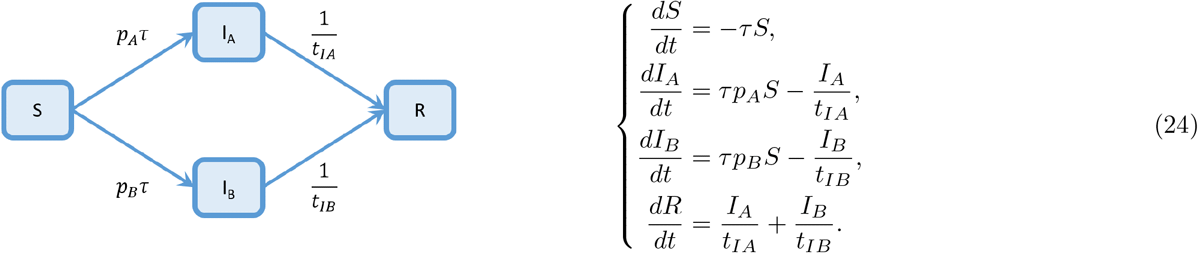

The fourth model considers two different sub-populations: say, females (compartments *S*_*f*_, *I*_*f*_ and *R*_*f*_) and males (compartments *S*_*m*_, *I*_*m*_ and *R*_*m*_):

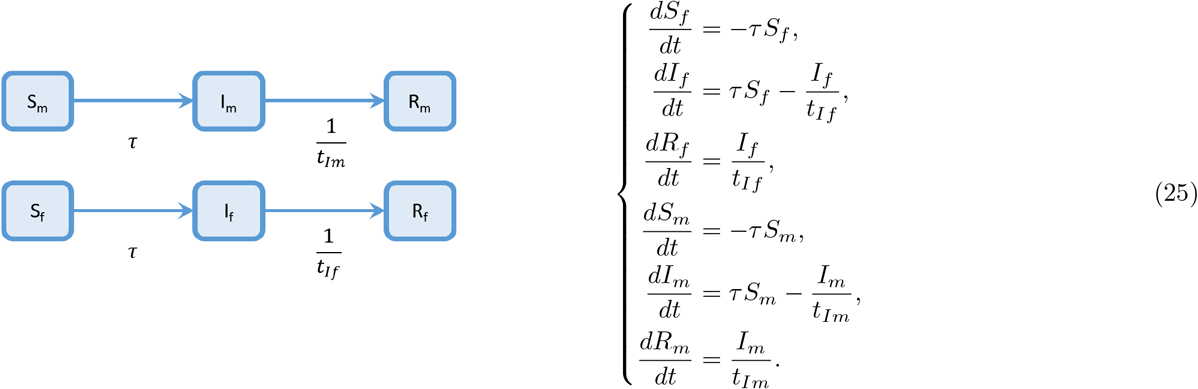

Let us assume that all compartments of all models are initially empty, except for *S, S*_*f*_ and *S*_*m*_ that are related at *t* = 0 by the constraint *S*_*f*0_ + *S*_*m*0_ = *S*_0_. In such a simple case, the analytical form of the total number of infected cases in the four models is given by the four equations:

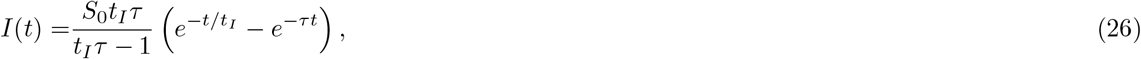

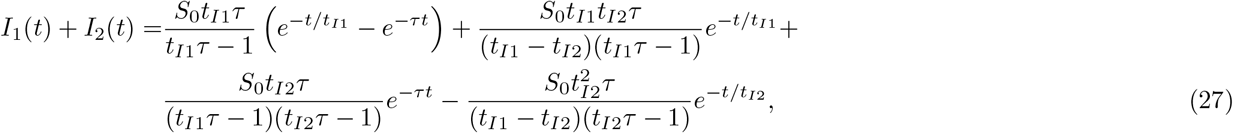

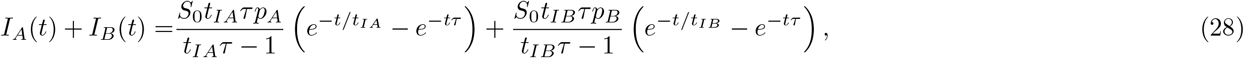

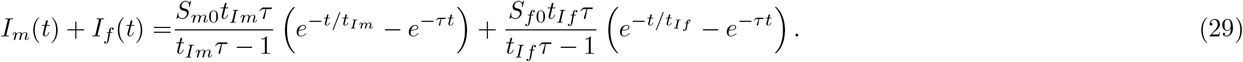

In particular cases, some approximations or equivalences can be obtained, as for instance:

- If *t*_*I*_ = *t*_*I*1_ + *t*_*I*2_ and *t*_*I*2_ is sufficiently small vs. *t*_*I*1_, then *I*(*t*) *≈ I*_1_(*t*) + *I*_2_(*t*);
- If *t*_*IA*_ = *t*_*IB*_ = *t*_*I*_ then *I*(*t*) = *I*_*A*_(*t*) + *I*_*B*_(*t*);
- If *t*_*If*_ = *t*_*Im*_ = *t*_*I*_ then *I*(*t*) = *I*_*f*_ (*t*) + *I*_*m*_(*t*).

However, in general, there is no direct identification of the parameters of the models 2, 3, and 4 to the parameters of the first model to have the same number of infected cases over time (i.e., *I*(*t*) = *I*_1_(*t*) + *I*_2_(*t*) = *I*_*A*_(*t*) + *I*_*B*_(*t*) = *I*_*f*_ (*t*) + *I*_*m*_(*t*)).

It is interesting to note that this comes from the non-stationarity of the system. Indeed, in an unrealistic, but stationary regime where the sizes of *S, S*_*f*_, *S*_*m*_ and all compartments of infected people are constant, then we have:

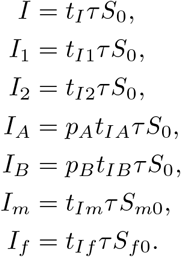

And the equivalence *I* = *I*_1_ + *I*_2_ = *I*_*A*_ + *I*_*B*_ = *I*_*f*_ + *I*_*m*_ can be ensured simply by choosing parameters that satisfy the constraints 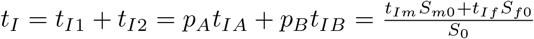. Of course if parameters satisfying these constraints are applied in a non-stationary case, the equivalence is no longer guarantied. As for the setting used Figure C.1, that shows substantial differences between models during transient regimes.

### C.2 Illustration with the modification of the compartments of SEIRAHm

As shown in Figure 3, the original SEIRAHm estimates much less individuals in compartment *I* than EHESPm in compartments *I* + *I*_*nh*_. One reason for this difference could be the presence of compartment *I*_*nh*_ in EHESPm. To test this hypothesis, a compartment *I*_*nh*_ was added to the SEIRAHm. In the resulting model, the average time spent in compartment *I* is *t*_*bh*_, and the average time spent in compartment *I*_*nh*_ is *t*_*s*_ − *t*_*bh*_ So, this part of the model becomes similar to the same part in EHESPm. Figure C.2 shows that this change has an important impact on the results; the number of individuals in compartment *I* in the modified SEIRAHm becomes much closer to the one estimated in EHESPm (when compared to Figure 3).

**Figure C.1:**
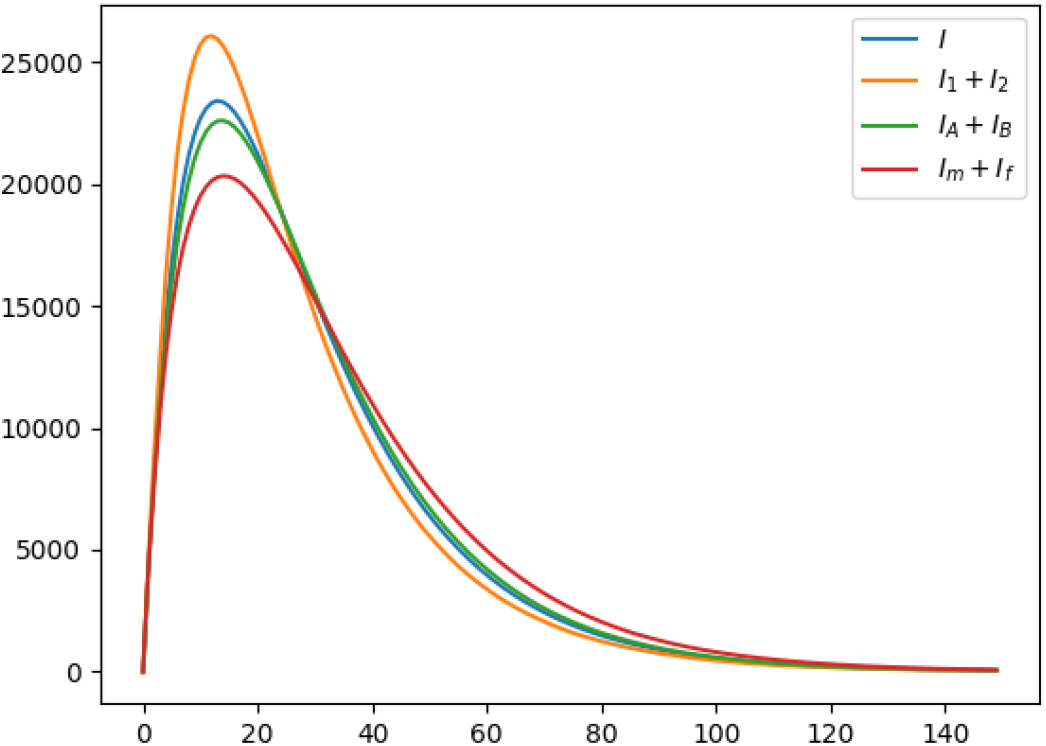
Dynamics of the total number of infected cases with each of the four models.

**Figure C.2:**
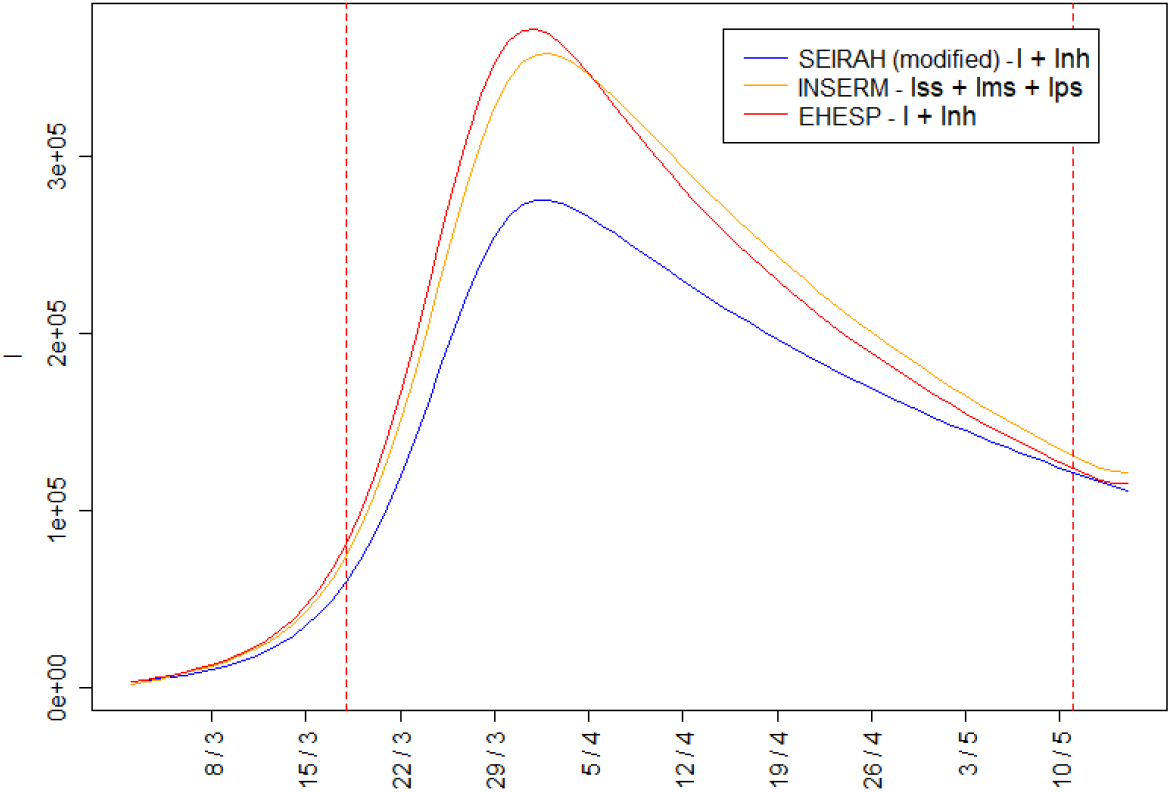
Number of individuals in compartments *I* over time in whole France (*I* + *I*_*nh*_ in EHESPm and in modified SEIRAHm, and *I*_*ps*_ + *I*_*ms*_ + *I*_*ss*_ in INSERMm).

### C.3 Other figures for compartment size comparison made Section 4.1

Output flow and variations of compartment I are given Figure C.3. Figure C.4 shows the comparison of the terms of Equation 9.

**Figure C.3:**
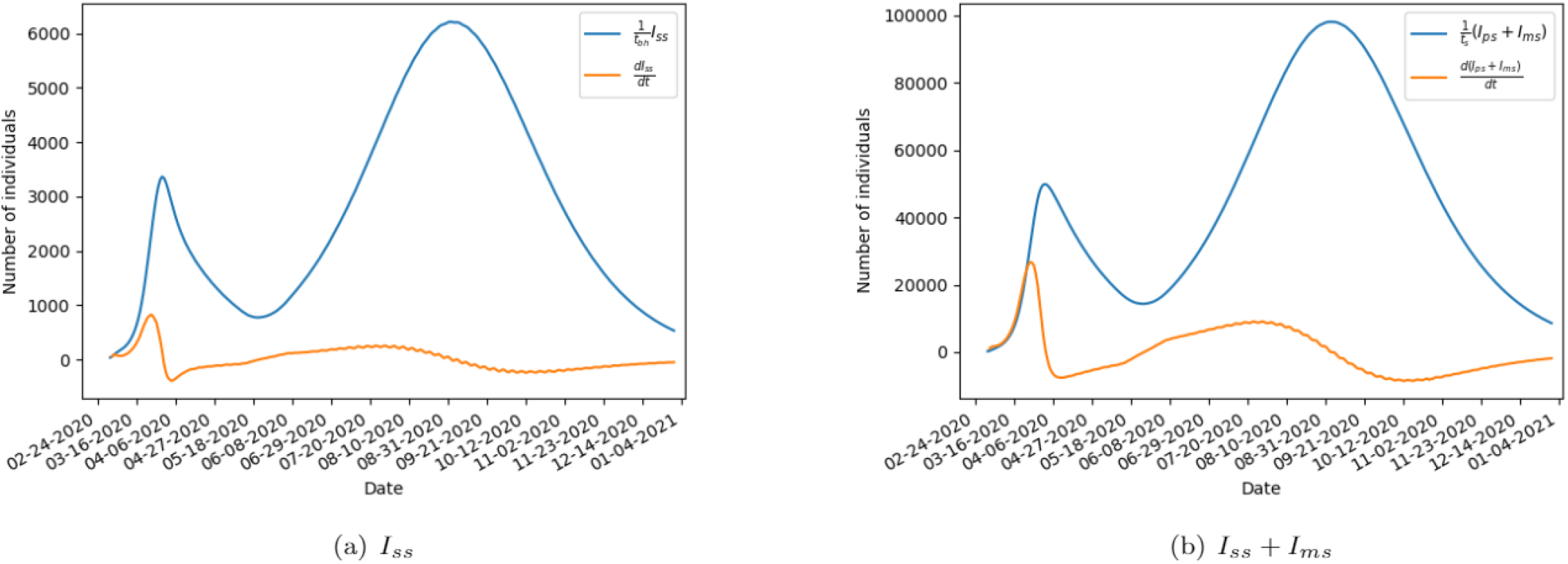
Output flow of compartment *I* and the variations of compartment *I* population for INSERMm for whole France, using *R*_0_ = 1.5 after first lockdown.

**Figure C.4:**
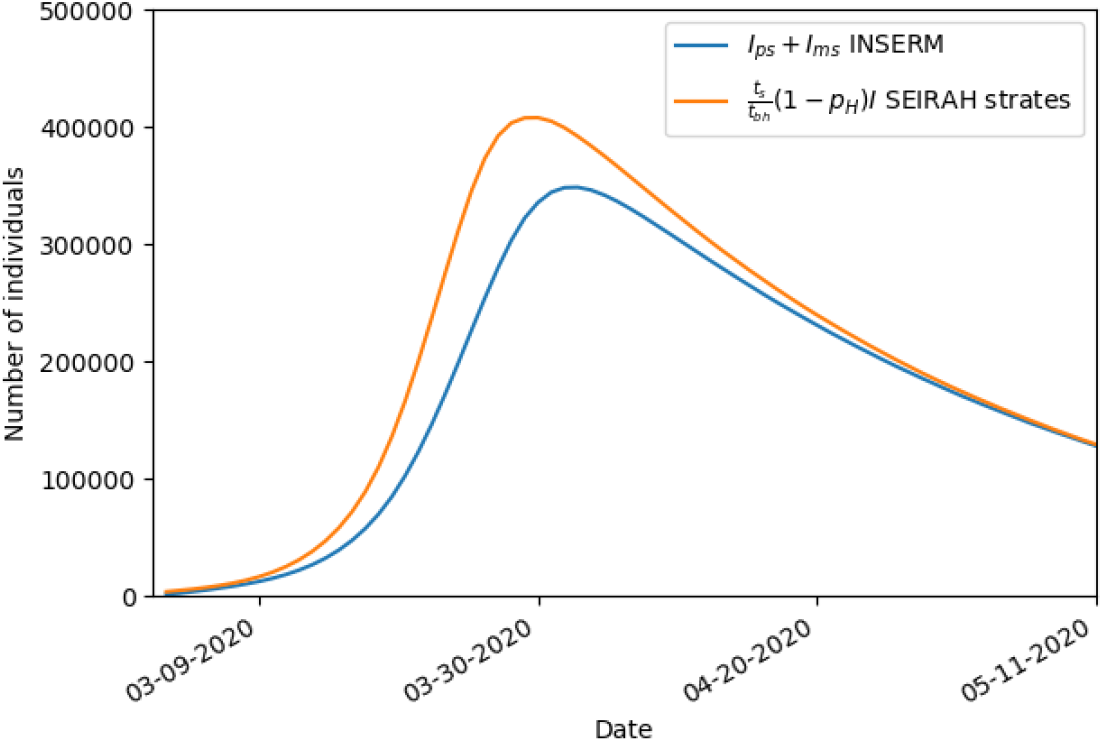
Comparison of the terms of Equation 9 for SEIRAHm and INSERMm compartments. To eliminate differences due to age classes, a SEIRAHm with age classes was used.

Members of the CovDyn (Covid Dynamics) group, by main affiliation

*LBBE – HCL* Mathieu FAUVERNIER, François GUEYFFIER, Jean IWAZ, Delphine MAUCORT-BOULCH, Simon PAGEAUD, Muriel RABILLOUD, Pascal ROY.

*LSAF* Alexis BIENVENÜE, Anne EYRAUD-LOISEL, Romain GAUCHON, Pierre-Olivier GOFFARD, Nico-las LEBOISNE, Stéphane LOISEL, Xavier MILHAUD, Denys POMMERET, Yahia SALHI, Pierre VANDEKERKHOVE.

*CIRI-HCL* Philippe VANHEMS.

*LMFA* Jean-Pierre BERTOGLIO.

*LTDS* Nicolas PONTHUS.

*INL* Christelle YEROMONAHOS.

*LIRIS* Stéphane DERRODE, Véronique DESLANDRES, Mohand-Saïd HACID, Salima HASSAS, Catherine POTHIER, Christophe RIGOTTI.

*ICJ* Thibault ESPINASSE, Samuel BERNARD, Philippe MICHEL, Vitaly VOLPERT.

*INRIA* Mostafa ADIMY.

*CHU De Rouen Unité INSERM 1018, CESP* Jacques BENICHOU.

*CHU De Rouen LIMICS INSERM U1142, Université de Rouen/ Sorbonne Université* Stéfan DARMONI.

*CHU de NICE, Université de Nice* Pascal STACCINI.

## Footnotes

[1] The lockdown began on March 17 at noon, which resulted in a large movement of populations in the morning. We assume thus that the lockdown started on March 18.

[2] The case of a sudden change of *β*_2,*t*_ at a time *Tc* was also tested. However, this did not change any of the conclusions of the paper.

[3] See 2.2.3 for the choice of, *T*_*p*_ and 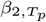.

[4] The short phase following contamination without pathological symptoms but with possible non-specific prodromes.

[5] The presence of a prodromic compartment implied that the average time during which an individual is infectious was higher in INSERMm than in SEIRAHm.

[6] This is true except at the beginning and end of the epidemic, Figures C.3.

[7] Figure C.4 shows the plots of the two terms of Equation 9 over the fitting period.

[8] Average of the values of *Tc* as estimated by F-WL approach and reported in Table B.1

[9] For the original authors of SEIRAHm [5], it was the starting date of the epidemic in many regions

[10] At a given date *t*, the value 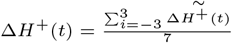 is retained, with 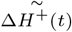 denoting the observed values.

[11] The compartments are ordered as follows : *E*,*I*_*p*_,*A,I*_*ps*_,*I*_*ms*_,*I*_*ss*_,*H* and *ICU*.

[12] The compartments are ordered as follows: *E,I*_*p*_,*A,I,I*_*nh*_ and *H*.

